# Mendelian randomization, lipids and coronary artery disease: trade-offs between study designs and assumptions

**DOI:** 10.1101/2024.10.01.24314568

**Authors:** Joy Shi, Sonja A. Swanson, Elizabeth W. Diemer, Hanna Gerlovin, Daniel C. Posner, Peter W.F. Wilson, J. Michael Gaziano, Kelly Cho, Miguel A. Hernan, the VA Million Veteran Program

**Affiliations:** VA Boston Healthcare System, Boston, Massachusetts; CAUSALab, Harvard T.H. Chan School of Public Health, Boston, Massachusetts; Department of Epidemiology, Harvard T.H. Chan School of Public Health, Boston, Massachusetts; Department of Epidemiology, University of Pittsburgh, Pittsburgh, Pennsylvania; Massachusetts Area Veterans Epidemiology Research and Information Center (MAVERIC), VA, Cooperative Studies Program, VA Boston Healthcare System, Boston, Massachusetts; Veterans Affairs Atlanta Healthcare System, Decatur, Georgia; Division of Cardiology, Emory University School of Medicine, Atlanta, Georgia; Department of Epidemiology, Rollins School of Public Health, Emory University, Atlanta, Georgia; Department of Medicine, Harvard Medical School, Boston, Massachusetts; Department of Medicine, Division of Aging, Brigham and Women’s Hospital, Boston, Massachusetts

**Keywords:** Mendelian randomization, instrumental variable, lipids, coronary artery disease

## Abstract

**Background:** Mendelian randomization (MR) studies have been described as naturally occurring randomized controlled trials (RCTs). However, MR often deviates from appropriate RCT design principles and relies heavily on two-sample approaches. We used data from the Million Veteran Program (MVP) to empirically evaluate the impact of study design choices and use of one-versus two-sample MR in a study of lipids and coronary artery disease.

**Methods:** Our MR study included MVP participants of European descent with no history of coronary artery disease or contraindications to low-density lipoprotein cholesterol (LDL-C)-related therapies. We sequentially modified the eligibility criteria, study duration and follow-up to reflect common study design decisions for MR. In all designs, we used one- and two-sample approaches to estimate 10-year risks of coronary artery disease per 39 mg/dL increase in LDL-C or 15.6 mg/dL increase in high-density lipoprotein cholesterol (HDL-C).

**Results:** For LDL-C, one-sample estimates varied across designs (odds ratios from 1.50 [95% CI: 1.34,1.68] to 2.23 [95% CI: 1.93,2.59]) and were most sensitive to the inclusion of prevalent outcome events in the analysis. Odds ratios obtained via two-sample MR were attenuated (1.13 [95% CI: 1.01,1.26] to 1.30 [95% CI: 1.15,1.46]). For HDL-C, we observed inverse or null relationships and estimates were qualitatively similar across all designs (odds ratios from 0.76 [95% CI: 0.68,0.86] to 0.93 [95% CI: 0.65,1.34]).

**Conclusions:** MR estimates can, in practice, be impacted by decisions in study design due to trade-offs between different biases, and investigators should evaluate the sensitivity of their estimates to different design decisions.

**Key Messages:**

- The analogy between Mendelian randomization (MR) studies and randomized trials is challenged by the use of two-sample approaches and deviations in study design that arise due to time zero misalignments in MR studies.
- MR estimates for the relationship between blood lipids and coronary heart disease in the Million Veteran Program were sensitive to certain study design decisions (such as whether prevalent outcome events were included in the analysis) and the use of one-sample versus two-sample approaches.
- Investigators should consider the sensitivity of their estimates to different MR designs and its implications for trade-offs between different sources of bias.

## Introduction

Mendelian randomization (MR) studies are observational studies that rely on instrumental variable estimation with genetic variants proposed as instruments to estimate causal effects. Within cardiovascular epidemiology, MR studies have concluded that low-density lipoprotein cholesterol (LDL-C) increases, and high-density lipoprotein cholesterol (HDL-C) has no effect on the risk of coronary artery disease.^1–5^ These MR results were compatible with findings from randomized trials that reported a beneficial effect of LDL-C lowering therapies (e.g., statins) and no effect or a harmful effect of HDL-C increasing therapies (e.g., cholesteryl ester transfer protein inhibitors).^6–11^

Per this example, MR studies have often been characterized as akin to randomized trials because genetic variants are randomly inherited at conception.^12–14^ However, this analogy is imperfect because MR estimates are more closely aligned with estimated effects under full adherence to the exposure (akin to per-protocol effects in randomized trials) rather than effects of assignment to the exposure (akin to intention-to-treat effects) and because the design of MR studies does not map cleanly to that of randomized trials.^15^

Because adherence-adjusted effects are the target, the validity of MR effect estimates requires that the genetic variant meets the instrumental conditions and other conditions required for point identification.^16–18^ Even more conditions are required in the many “two-sample” MR studies conducted by combining data on gene-exposure and gene-outcome relationships from two different data sources—analogous to combining the intention-to-treat estimate and the adherence estimate from separate trials.^15^ Furthermore, comparisons between MR studies and randomized trials are complicated by the fact that genetic variants are “assigned” at conception while the follow-up of MR studies typically starts decades after “assignment”.^15,19–21^ This design is akin to that of a randomized trial in which surviving participants are recruited long after being assigned to treatment.

The practical impact of violations of the instrumental conditions on MR estimates has been studied extensively.^22–25^ In contrast, few studies have assessed the impact of two-sample MR and other deviations from appropriate study design, partly because of the scarcity of large observational cohorts with genetic and longitudinal data collected over extended periods. Here, we used data from the Million Veteran Program (MVP), a large US biobank with linked medical records and questionnaire data, to empirically evaluate the impact of one-versus two-sample approaches and of different design choices for MR studies.

### Differences between the design of a randomized trial and an MR study

Consider a randomized trial of lipid-modifying therapies for the primary prevention of coronary artery disease. Eligible individuals (e.g., with no history of coronary heart disease) are enrolled, randomly assigned to a treatment strategy (e.g., statin therapy or no statin therapy), and followed from the time of assignment, which becomes time zero of follow-up (Figure 1i).

**Figure 1.**
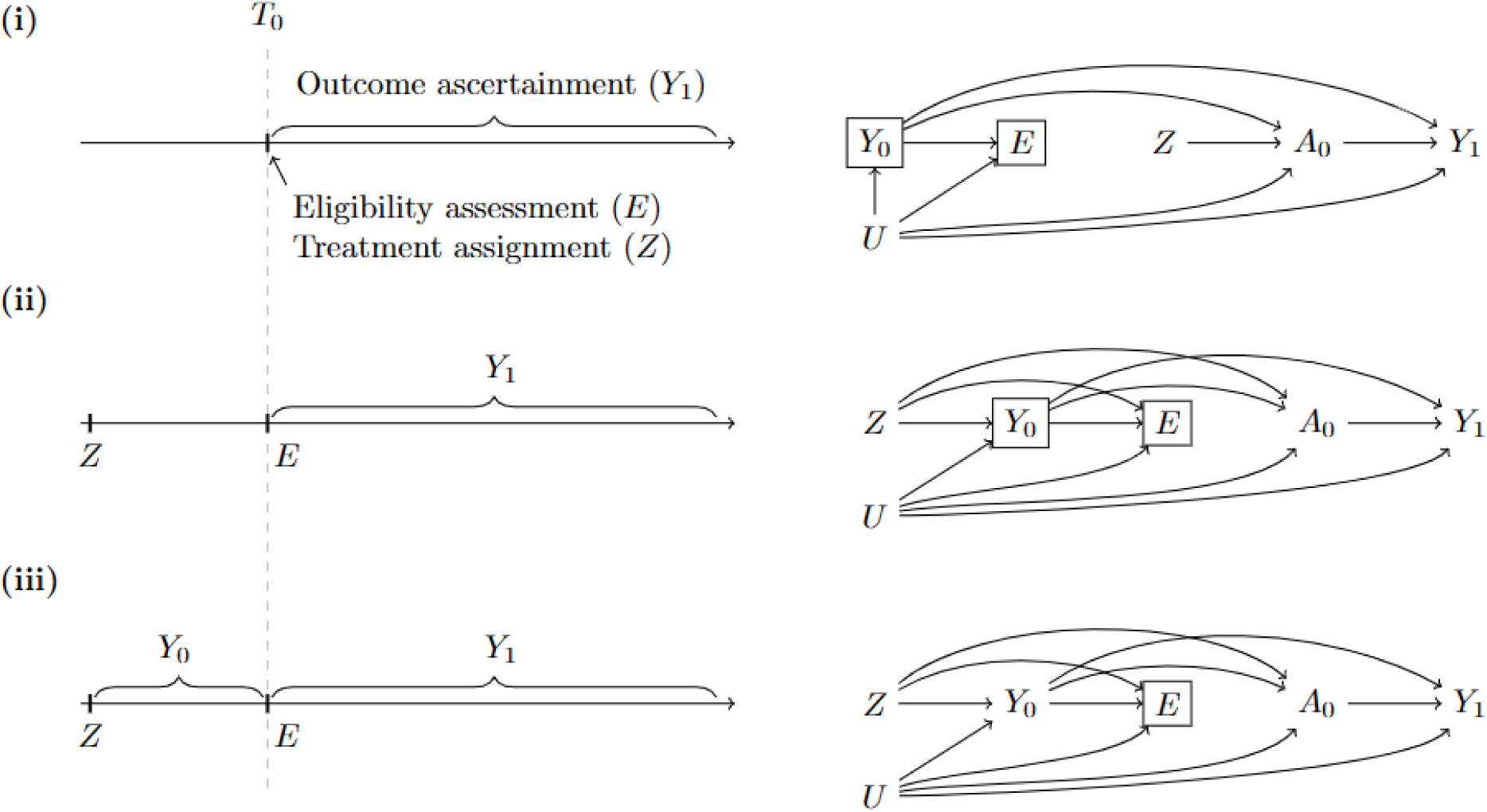
On the left, timing of eligibility assessment (*E*), treatment assignment (*Z*), and ascertainment of outcome events (*Y*_0_ and *Y*_1_) relative to time zero (*T*_0_) in (i) a randomized trial, (ii) a Mendelian randomization study with incident outcomes only, and (iii) a Mendelian randomization study with prevalent and incident outcomes. On the right, causal diagrams for each corresponding study design, represented by proposed instrument *Z* (randomized treatment assignment in a randomized trial; genetic variant(s) in a MR study), exposure *A*_0_, outcome status at end of follow-up *Y*_1_, outcome status at start of follow-up *Y*_0_, and eligibility at start of follow-up *E*.

In MR studies of coronary artery disease, individuals are “randomly assigned” genetic variants at conception, then typically enrolled and followed starting in middle or old age—decades after the “assignment” (Figure 1ii-iii).^15^ Therefore, genetic variants related to survival until the start of follow-up may spuriously associate with coronary artery disease, even if the variants have no effect on the outcome later in life.^26^ This selection bias is akin to the post-baseline selection bias that may arise in randomized trials with losses to follow-up.

Given the above, it is unclear how best to design MR studies. For example, a randomized trial for statins would exclude participants with a history of the outcome (i.e., restrict to participants with *Y*_0_ = 0 in Figure 1i). In MR studies, however, either including or excluding individuals with history of the outcome may introduce bias because (i) an MR study with only incident outcome events would be biased if the genetic variants are related to development of the outcome by the start of follow-up (Figure 1ii), and (ii) an MR study with both prevalent and incident outcome events may violate the exclusion restriction (via the path *Z* to *Y*_0_ to *Y*_1_ in Figure 1iii) and introduce bias due to reverse causation (where the effect of *Y*_0_ on *A*_0_ is misattributed to the effect of *A*_0_ on *Y*_1_).^27,28^ Decisions in MR study design present trade-offs between different biases.

### Differences between one-sample MR vs two-sample MR

MR requires estimates for both the gene-exposure and gene-outcome associations. These estimates may be obtained from a single data source (referred to as one-sample MR) or combined from two different data sources using meta-analytic approaches (referred to as two-sample MR). Note that, regardless of the choice of one-sample versus two-sample MR, a separate data source should be used for the selection of genetic variants proposed as instruments to avoid estimates being impacted by the “winner’s curse”.^29^

Two-sample MR studies are popular because of the growing availability of summary statistics from genetic consortia. It has also been argued that weak instruments bias estimates towards the null in two-sample MR but may bias estimates in either direction in one-sample MR.^30–32^ However, two-sample MR is more vulnerable to bias than one-sample MR because of possible heterogeneity in the distribution of confounders between the source populations, improper data harmonization, or inadequate analysis or study design that generated each of the summary statistics.^30,33,34^

### An MR study in the Million Veteran Program: Design

To illustrate the empirical impact of the decisions described above, we first outline the design of an MR study of LDL-C and HDL-C (our reference MR design) that deviates as minimally as possible from a randomized trial design. We then sequentially modify our design to reflect common decision points that investigators face when conducting MR. We start by describing the observational data.

#### Observational data

The Million Veteran Program (MVP) is an ongoing observational prospective cohort study and biobank of active users of the Veterans Health Administration (VHA) health care system.^35^ In brief, veterans were enrolled starting in 2011, and participation involved the completion of questionnaires, provision of a blood sample, and consent to data linkage with medical records and administrative health data. All participants signed informed consent, and the Veterans Affairs Central Institutional Review Board approved the study protocol.^35^ For this analysis, we used survey and electronic health record data from MVP core release v21.1, and genetic data from the MVP Release 4 dataset.^36^ Additional information on LDL-C and HDL-C measures and their proposed instruments, coronary artery disease events (both fatal and nonfatal, and included myocardial infarction, atherosclerosis, and undergoing angioplasty or revascularization), and other covariates are available in the Supplementary Methods and Table S1.

#### Our reference MR design

For this analysis, we included veterans who participated in MVP; were aged ≥18 years between January 1, 2002 and December 31, 2019; were of European descent; had known genotype information; and had known LDL-C or HDL-C in the past year. We considered only the most recent lipid measurement prior to the start of follow-up in our analyses. We excluded MVP participants with a history of the outcome (i.e., restricted to participants with *Y*_0_ = 0 in Figure 1ii) and with common contraindications for LDL-C lowering therapies (prior history of myopathy; chronic kidney disease or end-stage renal disease; chronic liver disease; and abnormal liver function).

For each eligible individual, we defined a single start of follow-up at the time in which all eligibility criteria were met (which may have occurred before or after MVP enrolment). Follow-up ended upon the earliest occurrence of the following: development of the outcome, loss to follow-up, 10 years after start of follow-up, or administrative end of follow-up (December 31, 2020). Note that selection bias can arise because veterans who meet the eligibility criteria at the start of follow-up will only be included in the analysis if they survive until MVP enrolment and agree to participate in MVP.

#### Modifications to the reference MR design

To evaluate the impact of different MR study design decisions on its estimates, we sequentially modified our reference design to emulate the design of previously published MR studies of lipids and coronary artery disease in the U.K. Biobank.^2,3^

1. Design variation 1: Eligible participants are restricted to age 40 to 69 years between January 1, 2006 and December 31, 2010 (based on the age and recruitment period for the U.K. Biobank).
2. Design variation 2: Analyses no longer adjust for loss to follow-up (i.e., censored participants are considered to have not developed the event), to evaluate the impact of ignoring losses to follow-up.
3. Design variation 3: Participants are followed until the earliest occurrence of the following: development of the outcome, death, or the administrative end of follow-up (March 31, 2017, previously December 31, 2020). Here, we assess the impact of evaluating outcome status at the end of administrative follow-up regardless of follow-up duration for a given individual.
4. Design variation 4: Participants are no longer excluded if they had a possible contraindication to a hypothetical LDL-C or HDL-C related therapy, to evaluate the impact of removing this eligibility criterion.
5. Design variation 5: Participants are no longer excluded if they had a prior history of coronary artery disease, to evaluate the impact of including prevalent outcome events in the analysis.

### An MR study in the Million Veteran Program: Data Analysis

We analysed the data under the reference design and each design variation using both one-sample and two-sample approaches, and compared the estimates obtained across different approaches to two previously published two-sample MR studies.^2,3^

For the one-sample MR analyses, we applied two-stage least-squares regression (via linear regression in the first stage and logistic regression in the second stage) to obtain 10-year odds ratios for coronary artery disease. All analyses adjusted for age at the start of follow-up, sex and ten genetic principal components of ancestry. To account for potential selection bias due to loss to follow-up (in the reference design and design variation 1), nonstabilized inverse probability of censoring weights were constructed using age at the start of follow-up, sex, ten genetic principal components, the proposed instrument and LDL-C or HDL-C measurement at the start of follow-up. We also considered analyses that restricted on age group (≤50, >50 to ≤60, >60 to ≤70) and on prior use of a known LDL-C lowering therapy (e.g., statins) in the 12 months before start of follow-up. Note that the same participant may have contributed to multiple age group-specific analyses if all other eligibility criteria were met at multiple ages.

For the two-sample MR analyses, inverse-variance weighting was used in the primary analysis, and MR-Egger regression and weighted median regression were conducted as sensitivity analyses. Gene-lipid associations were obtained from the Global Lipids Genetics Consortium (GLGC),^37^ and gene-outcome associations (with adjustment for age at the start of follow-up, sex and ten genetic principal components) were estimated in MVP. Modifications to the reference design were implemented in MVP only where individual-level data were available. Age- and LDL-C lowering therapy use-restricted analyses (for the gene-outcome associations) were conducted as described above.

## Results

The cohort included 366,782 eligible participants from the Million Veteran Program, of whom 365,725 were included in analyses of LDL-C and 366,515 in analyses of HDL-C (Figure 2). At the time of first eligibility, their average age was 55 years, 91% were male, and 13% had used an LDL-C lowering therapy in the previous year (Table 1).

**Figure 2.**
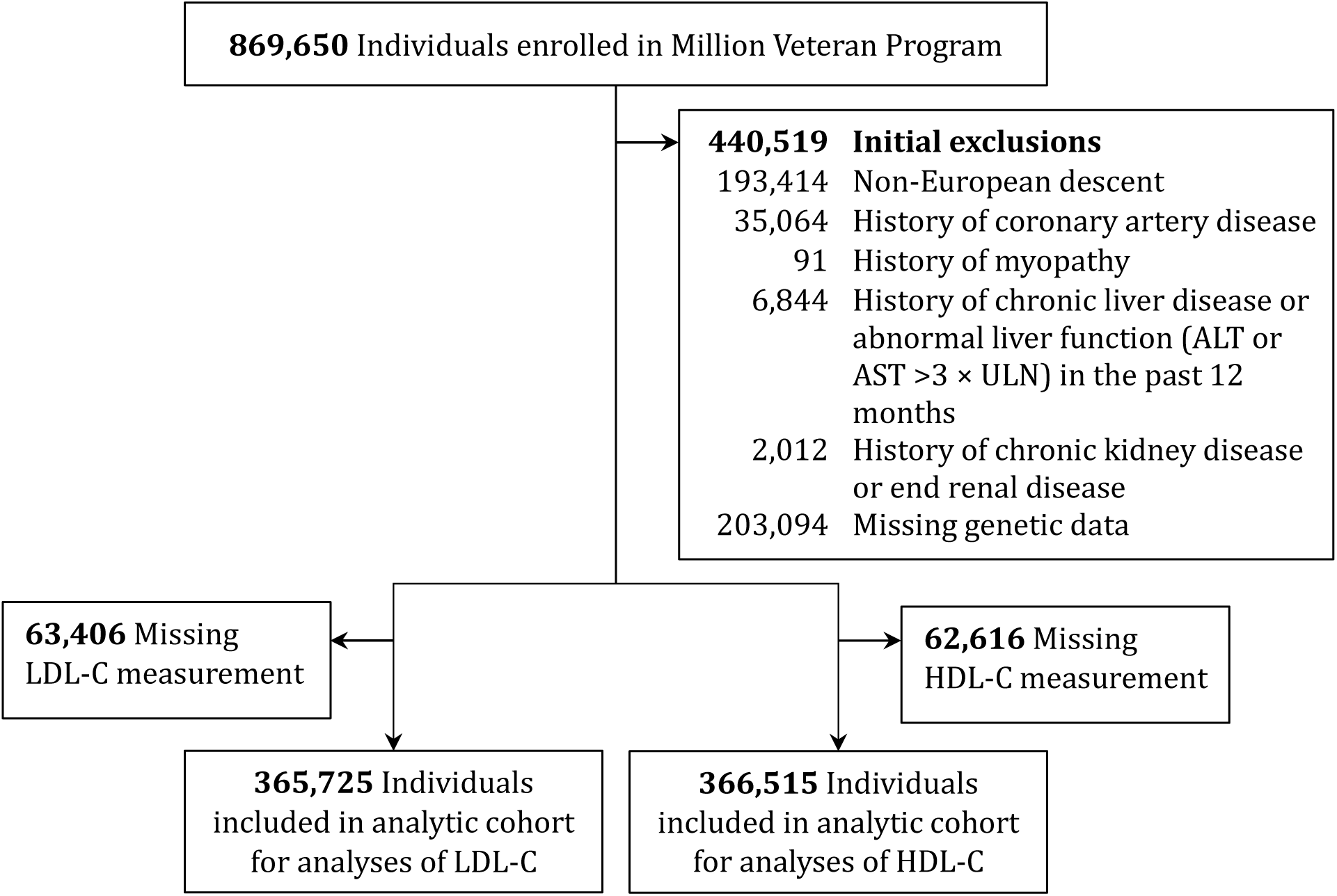
Selection of eligible individuals for an MR analysis of LDL-C, HDL-C and coronary artery disease using data from the Million Veteran Program.

**Table 1.**
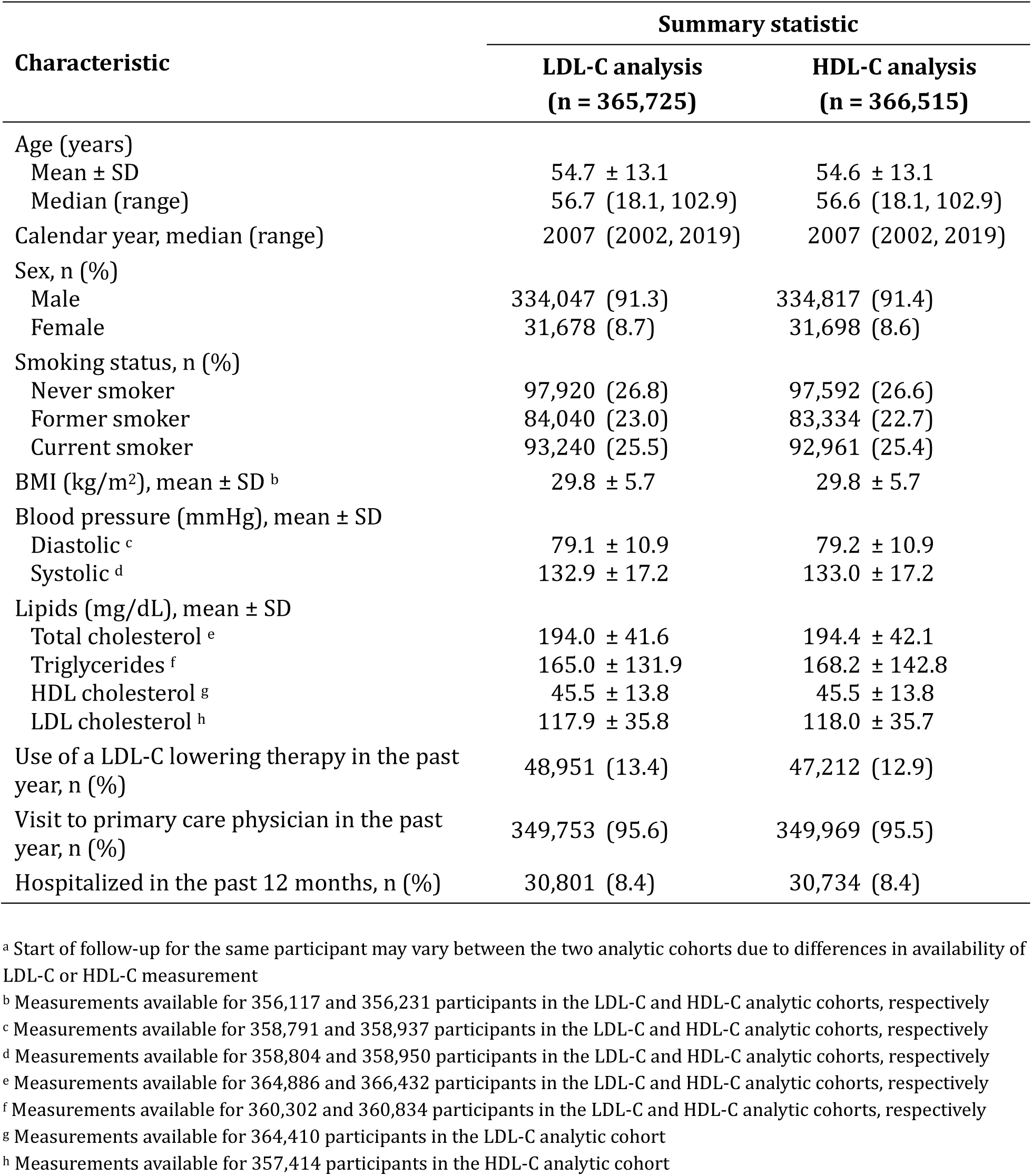
Characteristics at time of first eligibility for MVP participants included in MR analyses of LDL-C and HDL-C ^a^.

Figure 3 presents the odds ratios for coronary artery disease that correspond to a 39 mg/dL (or 1 mmol/L) increase in LDL-C. In our reference MR design, the 10-year odds ratio was 1.50 (95% CI: 1.34,1.68) (Table S4). With each sequential modification to the reference MR design, the estimates obtained via a one-sample approach incrementally shifted further from the null and ranged from 1.61 (95% CI: 1.35,1.86) to 2.23 (95% CI: 1.93,2.59). The estimates obtained via a two-sample approach were smaller in magnitude, ranging from 1.13 (95% CI: 1.01,1.26) to 1.30 (95% CI: 1.15,1.46) when inverse-variance weighting was applied. Sensitivity analyses using MR-Egger regression and weighted median regression resulted in similar estimates (Table S4). In age-restricted analyses, estimates in the ≤50 age group were closer to the null across all designs although confidence intervals were wide (Figure 4, Table S5). In analyses among participants who had no prior use of a known LDL-C lowering therapy in the 12 months before the start of follow-up, estimates were shifted downwards relative to estimates in the overall population (Figure S1). The shift in estimates was larger with increasing modifications to the reference MR design, and in older age groups where exclusions were greater (32% to 58% of these participants were excluded due to the use of LDL-C lowering therapy at the start of follow-up) (Table S5).

**Figure 3.**
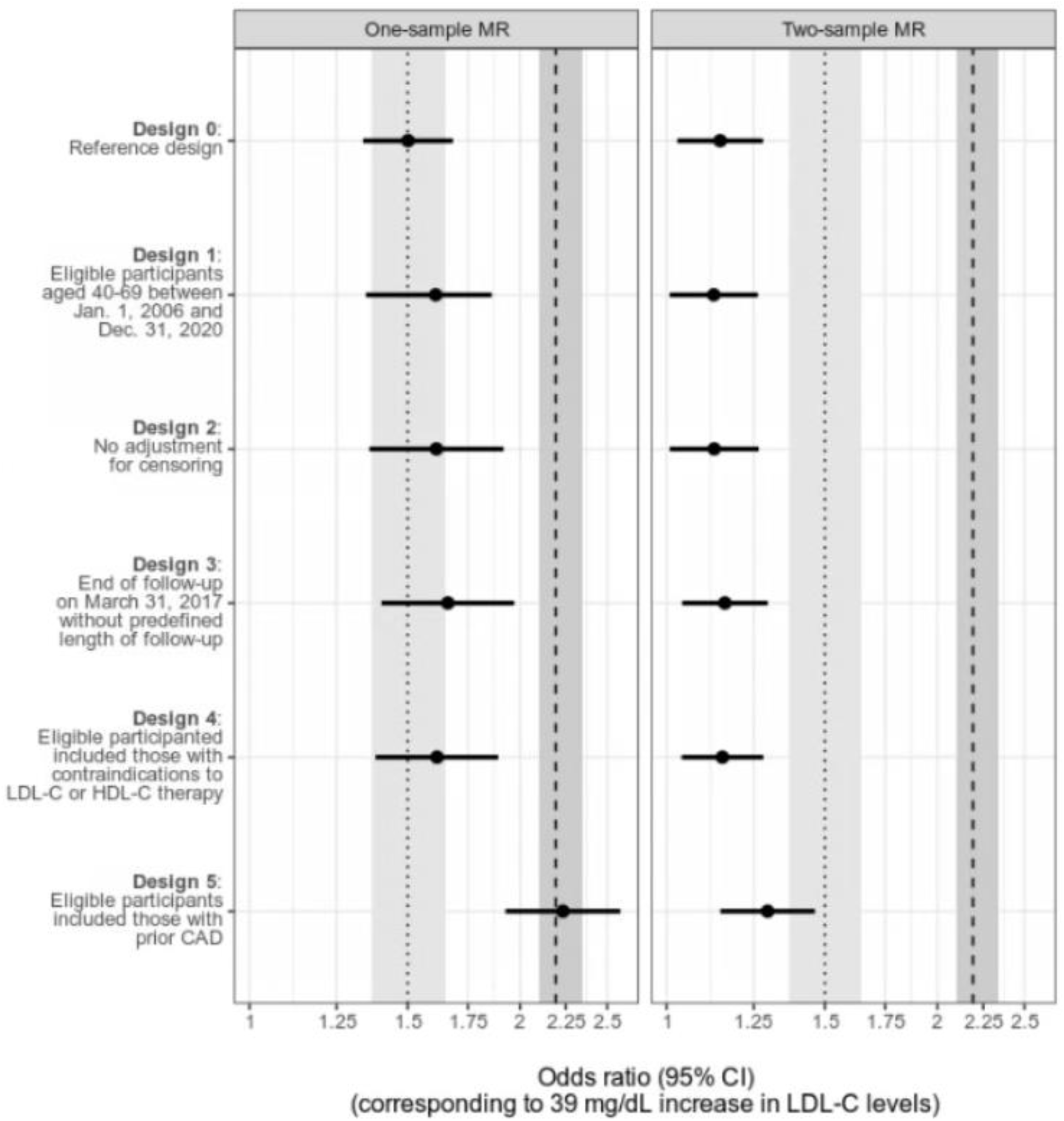
MR estimates of odds ratios and 95% confidence intervals for LDL-C, Million Veteran Program. The dotted and dashed lines and shaded light and dark grey areas correspond to the point estimates and 95% confidence intervals obtained from representative two-sample MR studies of designs 4 and 5, respectively (Allara et al., 2019; Ferrence et al., 2019)

**Figure 4.**
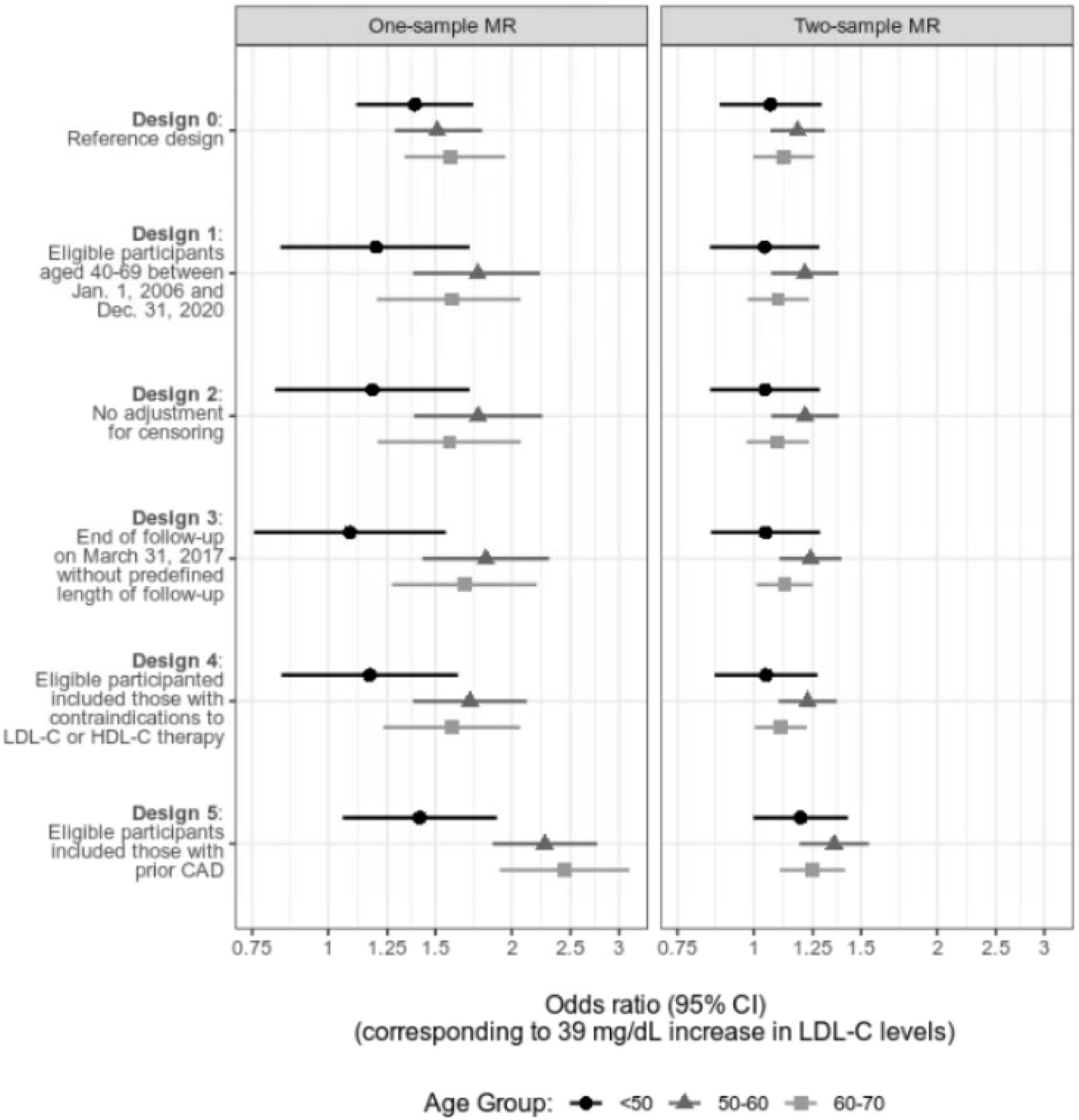
MR estimates of odds ratios and 95% confidence intervals for LDL-C, across different age groups (<50, 50 to <60, 60 to <70), Million Veteran Program.

In Figure 5, odds ratios for coronary artery disease that correspond to a 15.6 mg/dL increase in HDL-C are presented. We observed a 10-year odds ratio of 0.76 (95% CI: 0.68,0.86) in our reference MR design (Table S6). Subsequent modifications to the study design resulted in one-sample estimates that were shifted upwards relative to the estimate obtained from our reference design, ranging from 0.78 (95% CI: 0.58,1.07) to 0.93 (95% CI: 0.65,1.34). Our one-sample estimates were similar in magnitude to the two-sample estimates obtained via inverse-variance weighting, whereas the two-sample estimates obtained via MR-Egger regression and weighted median regression were closer to the null (Table S6). Similar estimates were also obtained in age-restricted analyses (Figure S2). Sensitivity analyses based on an alternate definition of coronary artery disease resulted in similar estimates (Table S7).

**Figure 5.**
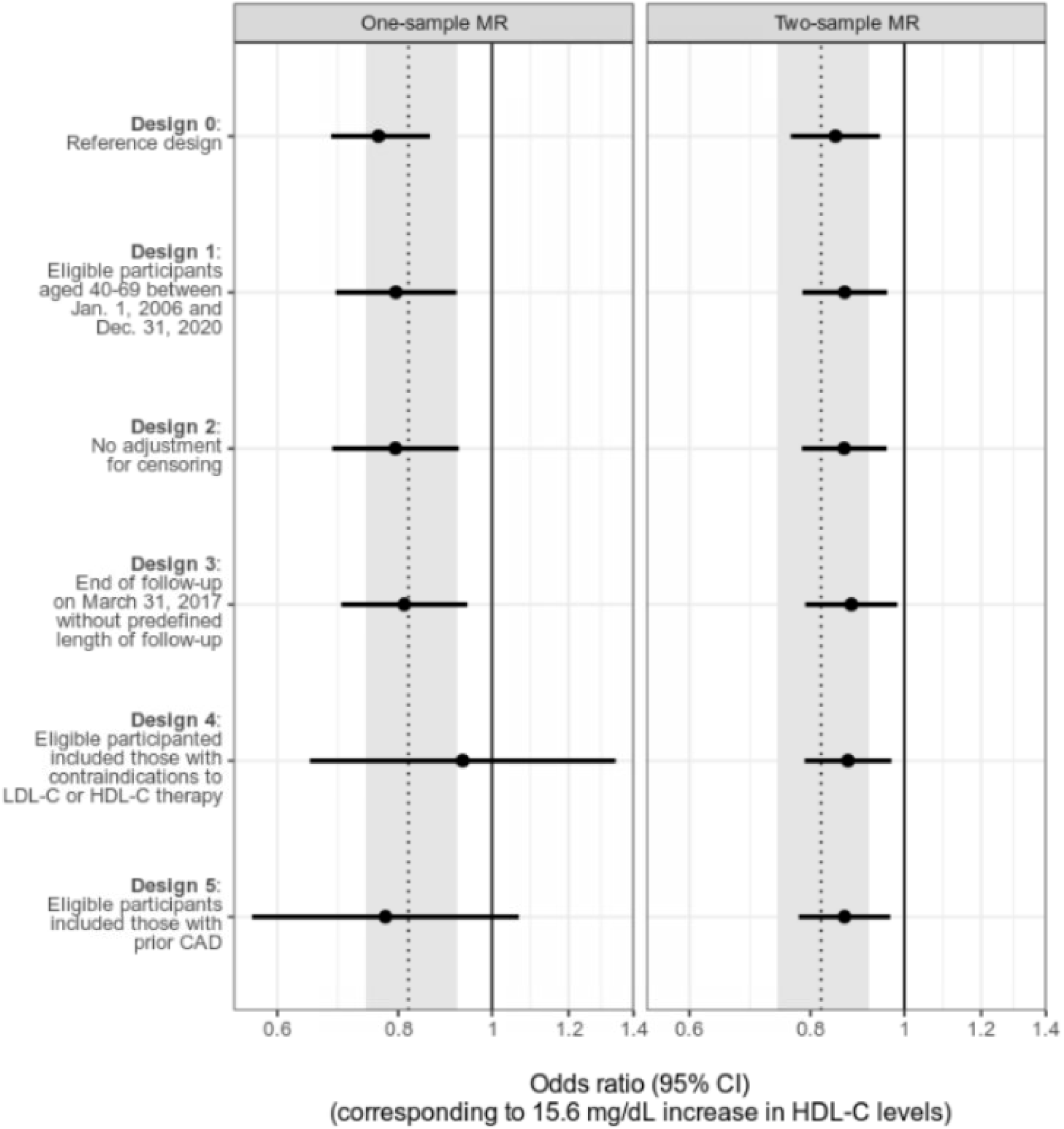
MR estimates of odds ratios and 95% confidence intervals for in HDL-C, Million Veteran Program. The dotted line and shaded light grey area correspond to the point estimate and 95% confidence interval obtained a representative two-sample MR study of design 4 (Allara et al., 2019)

## Discussion

We used MR to evaluate the relationships of LDL-C and HDL-C with coronary artery disease in the Million Veteran Program. Our estimates were qualitatively consistent with previous findings of a positive association between LDL-C and coronary artery disease risk, and a null or inverse association between HDL-C and coronary heart disease risk.^2,3^ However, the magnitude of our estimates was heterogeneous across different study designs and analytical approaches.

Despite frequent comparisons between MR and randomized trials, MR studies often deviate in key study design features. For LDL-C, our one-sample MR estimate was not sensitive to changes in follow-up (between design variation 1 and 3) or eligibility criteria (between design variation 3 and 4). This is likely due to minimal losses to follow-up in our population and suggests that these additional eligibility criteria did not introduce bias. However, we observed a large change in our estimate between design variations 4 and 5 (from an odds ratio of 1.62 to 2.23) when prevalent outcome events were included in the analysis, suggesting there is a large trade-off in biases (as described above) when including versus excluding prevalent outcomes. These estimates (from design variations 4 and 5) were compatible with estimates from previous studies (Allara and colleagues, and Ference and colleagues, respectively).^2,3^ For HDL-C, we observed inverse or null relationships with coronary artery disease and estimates were qualitatively similar across all designs. This is also consistent with findings from Allara and colleagues, who reported inverse associations in multivariable and univariable analyses using inverse-variance weighting, but were null in univariable analyses using MR-Egger regression.^2^

Our two-sample MR estimates for LDL-C were more attenuated, which may be explained by differences in the source populations for GLGC and MVP. GLGC is 52% male and excludes participants known to be on lipid-lowering medication, whereas MVP is 90% male and composed entirely of military veterans whose use of lipid-lowering medications is high. A second explanation is that our two-sample MR estimates were influenced by the winner’s curse since GLGC was used for both variant selection and estimation,^29^ although this approach is consistent with previous MR studies.^2^ Compared to our one-sample estimates, our two-sample estimates were also less sensitive to changes in the study design. One explanation is that changes to the study design could only be implemented in MVP because individual-level data were not available for GLGC. Of note, GLGC includes participants with prevalent outcome events and pooled data across 60 studies—some of which are cross-sectional in nature—resulting in an ill-defined follow-up period in the MR analysis.^37^

In age group-specific analyses, estimates were attenuated in the <50 age group, regardless of design or use of a one-versus two-sample approach, although confidence intervals were wide. This may suggest that the contributions of LDL-C and HDL-C to coronary artery disease risk may vary over time but could also be explained by increasing selection bias as age at the start of follow-up increases. For LDL-C, we also conducted analyses that were restricted to non-users of LDL-C lowering therapies in the year prior to start of follow-up—a common approach for handling medication use in MR studies of LDL-C. Although we observed some differences in the subgroup of non-users, this could be explained by potential selection bias introduced by restricting or stratifying on medication use.^20^

Our MR study was limited in its ability to precisely emulate certain study design features of randomized trials. First, as described above, time zero is misaligned in MR studies due to delays in the start of follow-up relative to conception.^15^ Second, the treatment strategies of interest are often ill-defined in MR studies. Changes in LDL-C can result from multiple versions of treatment (e.g., diet, different classes of LDL-C lowering therapies) that have varying effects on coronary artery disease, even if targeted to the same change in LDL-C levels.^38–41^ Third, our analysis would ideally incorporate longitudinal measurements of LDL-C and HDL-C over follow-up with time-to-event data on coronary artery disease. Although IV methods for time-varying exposures and time-to-event outcomes have been previously proposed,^42^ we applied conventional approaches for consistency and comparison with previous MR estimates, which can be interpreted as a “lifetime effect” assuming the gene-exposure relationship stays constant over time.^17,18^

In conclusion, the analogy between Mendelian randomization studies and randomized trials is challenged by the use of two-sample approaches and deviations from randomized trial study design. In MR studies, the delay between randomization and start of follow-up introduces different decisions that have trade-offs with respect to bias. We demonstrate the practical implications of these trade-offs in an analysis of lipids and coronary heart disease using data from the Million Veteran Program, although the potential bias introduced by different decisions are likely to be question-, context- and data-specific. Although our one-sample analyses using individual-level data from the Million Veteran Program were qualitatively compatible with findings from previous two-sample MR studies, estimates varied depending on whether prevalent outcome events were included in the analysis. Therefore, investigators should consider the sensitivity of their estimates to different MR designs and its implications for trade-offs between different sources of bias.

## Ethics approval

This study was conducted according to the guidelines laid down in the Declaration of Helsinki. The VA Central Institutional Review Board, Washington, DC, approved all study activities (protocol: MVP000, date of approval: 2010) and written informed consent was obtained from all participants.

## Data availability

MVP genomic and phenotypic data are currently only available to VA researchers through a research merit review process with VA’s Office of Research and Development (ORD).

## Author contributions

J.S., S.A.S., E.W.D. and M.A.H. conceptualized the study. H.G., D.C.P., K.C., J.M.G. and P.W.F.W. were involved in managing the project and curating the data. J.S. conducted the analysis and prepared the original manuscript draft. All authors have read and agreed to the final version of the manuscript.

## Acknowledgement

This research was supported by the Million Veteran Program (MVP#001) and Cooperative Studies Program (CSP#2032) from the Office of Research and Development, Veterans Health Administration, by resources and the use of facilities at the VA Boston Healthcare System and by the resources provided by the VA Informatics and Computing Infrastructure (VINCI) (VA HSR RES 13-457). Support for VA/CMS data provided by the Department of Veterans Affairs, VA Health Services Research and Development Service, VA Information Resource Center (Project Numbers SDR 02-237 and 98-004). J.S., S.A.S., E.W.D., H.G., and M.A.H. are supported by CSP #2032. The authors thank Juan P. Casas, the VA Informatics and Computing Infrastructure (VINCI) and Genomic Information System for Integrative Science (GenISIS) support teams and the MVP Core Statistical Analysis team for their contributions to this study, as well as the veterans who agreed to enrol in MVP. This publication does not represent the views of the Department of Veteran Affairs or the United States Government.

## References

1. Holmes MV, Asselbergs FW, Palmer TM, et al. Mendelian randomization of blood lipids for coronary heart disease. European Heart Journal. 2015;36(9):539–550. doi:10.1093/eurheartj/eht571

2. Allara E, Morani G, Carter P, et al. Genetic Determinants of Lipids and Cardiovascular Disease Outcomes: A Wide-Angled Mendelian Randomization Investigation. Circ: Genomic and Precision Medicine. 2019;12(12):e002711. doi:10.1161/CIRCGEN.119.002711

3. Ference BA, Bhatt DL, Catapano AL, et al. Association of Genetic Variants Related to Combined Exposure to Lower Low-Density Lipoproteins and Lower Systolic Blood Pressure With Lifetime Risk of Cardiovascular Disease. JAMA. 2019;322(14):1381. doi:10.1001/jama.2019.14120

4. Kawashiri M aki, Tada H, Nomura A, Yamagishi M. Mendelian randomization: Its impact on cardiovascular disease. Journal of Cardiology. 2018;72(4):307–313. doi:10.1016/j.jjcc.2018.04.007

5. Larsson SC, Butterworth AS, Burgess S. Mendelian randomization for cardiovascular diseases: principles and applications. European Heart Journal. 2023;44(47):4913–4924. doi:10.1093/eurheartj/ehad736

6. Cholesterol Treatment Trialists’ (Ctt) Collaborators. The effects of lowering LDL cholesterol with statin therapy in people at low risk of vascular disease: meta-analysis of individual data from 27 randomised trials. The Lancet. 2012;380(9841):581–590. doi:10.1016/S0140-6736(12)60367-5

7. Cholesterol Treatment Trialists’ (Ctt) Collaboration. Efficacy and safety of more intensive lowering of LDL cholesterol: a meta-analysis of data from 170 000 participants in 26 randomised trials. The Lancet. 2010;376(9753):1670–1681. doi:10.1016/S0140-6736(10)61350-5

8. Barter PJ, Caulfield M, Eriksson M, et al. Effects of Torcetrapib in Patients at High Risk for Coronary Events. N Engl J Med. 2007;357(21):2109–2122. doi:10.1056/NEJMoa0706628

9. The AIM-HIGH Investigators. Niacin in Patients with Low HDL Cholesterol Levels Receiving Intensive Statin Therapy. N Engl J Med. 2011;365(24):2255–2267. doi:10.1056/NEJMoa1107579

10. The HPS2-THRIVE Collaborative Group. Effects of Extended-Release Niacin with Laropiprant in High-Risk Patients. N Engl J Med. 2014;371(3):203–212. doi:10.1056/NEJMoa1300955

11. Lincoff AM, Nicholls SJ, Riesmeyer JS, et al. Evacetrapib and Cardiovascular Outcomes in High-Risk Vascular Disease. N Engl J Med. 2017;376(20):1933–1942. doi:10.1056/NEJMoa1609581

12. Sekula P, Del Greco M F, Pattaro C, Kottgen A. Mendelian Randomization as an Approach to Assess Causality Using Observational Data. J Am Soc Nephrol. 2016;27(11):3253–3265. doi:10.1681/ASN.2016010098

13. Roberts R. Mendelian Randomization Studies Promise to Shorten the Journey to FDA Approval. JACC: Basic to Translational Science. 2018;3(5):690–703. doi:10.1016/j.jacbts.2018.08.001

14. Ference BA, Holmes MV, Smith GD. Using Mendelian Randomization to Improve the Design of Randomized Trials. Cold Spring Harb Perspect Med. 2021;11(7):a040980. doi:10.1101/cshperspect.a040980

15. Swanson SA, Tiemeier H, Ikram MA, Herna n MA. Nature as a Trialist?: Deconstructing the Analogy Between Mendelian Randomization and Randomized Trials. Epidemiology. 2017;28(5):653–659. doi:10.1097/EDE.0000000000000699

16. Miguel HA, Robins JM. Causal Inference: What If. Chapman & Hall/CRC; 2021.

17. Labrecque JA, Swanson SA. Interpretation and Potential Biases of Mendelian Randomization Estimates With Time-Varying Exposures. Am J Epidemiol. 2019;188(1):231–238. doi:10.1093/aje/kwy204

18. Shi J, Swanson SA, Kraft P, Rosner B, De Vivo I, Herna n MA. Mendelian Randomization With Repeated Measures of a Time-varying Exposure: An Application of Structural Mean Models. Epidemiology. 2022;33(1):84–94. doi:10.1097/EDE.0000000000001417

19. Herna n MA, Sauer BC, Hernandez-Díaz S, Platt R, Shrier I. Specifying a target trial prevents immortal time bias and other self-inflicted injuries in observational analyses. Journal of Clinical Epidemiology. 2016;79:70–75. doi:10.1016/j.jclinepi.2016.04.014

20. Swanson SA. A Practical Guide to Selection Bias in Instrumental Variable Analyses. Epidemiology. 2019;30(3):345–349. doi:10.1097/EDE.0000000000000973

21. Vansteelandt S, Dukes O, Martinussen T. Survivor bias in Mendelian randomization analysis. Biostatistics. 2018;19(4):426–443. doi:10.1093/biostatistics/kxx050

22. Burgess S, Thompson SG, CRP CHD Genetics Collaboration. Avoiding bias from weak instruments in Mendelian randomization studies. Int J Epidemiol. 2011;40(3):755–764. doi:10.1093/ije/dyr036

23. Bowden J, Davey Smith G, Burgess S. Mendelian randomization with invalid instruments: effect estimation and bias detection through Egger regression. International Journal of Epidemiology. 2015;44(2):512–525. doi:10.1093/ije/dyv080

24. Jackson JW, Swanson SA. Toward a clearer portrayal of confounding bias in instrumental variable applications. Epidemiology. 2015;26(4):498–504. doi:10.1097/EDE.0000000000000287

25. Labrecque J, Swanson SA. Understanding the Assumptions Underlying Instrumental Variable Analyses: a Brief Review of Falsification Strategies and Related Tools. Curr Epidemiol Rep. 2018;5(3):214–220. doi:10.1007/s40471-018-0152-1

26. Herna n MA, Herna ndez-Dí az S, Robins JM. A structural approach to selection bias. Epidemiology. 2004;15(5):615–625. doi:10.1097/01.ede.0000135174.63482.43

27. Burgess S, Swanson SA, Labrecque JA. Are Mendelian randomization investigations immune from bias due to reverse causation? Eur J Epidemiol. 2021;36(3):253–257. doi:10.1007/s10654-021-00726-8

28. VanderWeele TJ, Tchetgen Tchetgen EJ, Cornelis M, Kraft P. Methodological challenges in Mendelian randomization. Epidemiology. 2014;25(3):427–435. doi:10.1097/EDE.0000000000000081

29. Taylor AE, Davies NM, Ware JJ, VanderWeele T, Smith GD, Munafo MR. Mendelian randomization in health research: Using appropriate genetic variants and avoiding biased estimates. Economics & Human Biology. 2014;13:99–106. doi:10.1016/j.ehb.2013.12.002

30. Hartwig FP, Davies NM, Hemani G, Davey Smith G. Two-sample Mendelian randomization: avoiding the downsides of a powerful, widely applicable but potentially fallible technique. International Journal of Epidemiology. 2016;45(6):1717–1726. doi:10.1093/ije/dyx028

31. Pierce BL, Burgess S. Efficient Design for Mendelian Randomization Studies: Subsample and 2-Sample Instrumental Variable Estimators. American Journal of Epidemiology. 2013;178(7):1177–1184. doi:10.1093/aje/kwt084

32. Davey Smith G, Hemani G. Mendelian randomization: genetic anchors for causal inference in epidemiological studies. Hum Mol Genet. 2014;23(R1):R89–98. doi:10.1093/hmg/ddu328

33. Zhao Q, Wang J, Spiller W, Bowden J, Small DS. Two-Sample Instrumental Variable Analyses Using Heterogeneous Samples. Statist Sci. 2019;34(2). doi:10.1214/18-STS692

34. Hartwig FP, Tilling K, Davey Smith G, Lawlor DA, Borges MC. Bias in two-sample Mendelian randomization when using heritable covariable-adjusted summary associations. Int J Epidemiol. 2021;50(5):1639–1650. doi:10.1093/ije/dyaa266

35. Gaziano JM, Concato J, Brophy M, et al. Million Veteran Program: A mega-biobank to study genetic influences on health and disease. J Clin Epidemiol. 2016;70:214–223. doi:10.1016/j.jclinepi.2015.09.016

36. Hunter-Zinck H, Shi Y, Li M, et al. Genotyping Array Design and Data Quality Control in the Million Veteran Program. Am J Hum Genet. 2020;106(4):535–548. doi:10.1016/j.ajhg.2020.03.004

37. Global Lipids Genetics Consortium. Discovery and refinement of loci associated with lipid levels. Nat Genet. 2013;45(11):1274–1283. doi:10.1038/ng.2797

38. Herna n MA. Invited Commentary: Hypothetical Interventions to Define Causal Effects— Afterthought or Prerequisite? American Journal of Epidemiology. 2005;162(7):618–620. doi:10.1093/aje/kwi255

39. Herna n MA. Does water kill? A call for less casual causal inferences. Annals of Epidemiology. 2016;26(10):674–680. doi:10.1016/j.annepidem.2016.08.016

40. Herna n MA, Taubman SL. Does obesity shorten life? The importance of well-defined interventions to answer causal questions. Int J Obes. 2008;32(S3):S8–S14. doi:10.1038/ijo.2008.82

41. VanderWeele TJ, Hernan MA. Causal inference under multiple versions of treatment. Journal of Causal Inference. 2013;1(1):1–20. doi:10.1515/jci-2012-0002

42. Shi J, Swanson SA, Kraft P, Rosner B, De Vivo I, Herna n MA. Instrumental variable estimation for a time-varying treatment and a time-to-event outcome via structural nested cumulative failure time models. BMC Med Res Methodol. 2021;21(1):258. doi:10.1186/s12874-021-01449-w

