## Supplemental Materials for "Mendelian randomization, lipids and coronary artery disease: trade-offs between study designs and assumptions"

**Supplementary Materials**

Joy Shi, Sonja A. Swanson, Elizabeth W. Diemer, Hanna Gerlovin, Daniel C. Posner, Kelly Cho, J. Michael Gaziano, Peter W.F. Wilson, Miguel A. Hernán, on behalf of the VA Million Veteran Program

### Acknowledgement List

**VA Million Veteran Program:**

**MVP Program Office**

- Program Director - Sumitra Muralidhar, Ph.D.
  US Department of Veterans Affairs, 810 Vermont Avenue NW, Washington, DC 20420
- Associate Director, Scientific Programs - Jennifer Moser, Ph.D.
  US Department of Veterans Affairs, 810 Vermont Avenue NW, Washington, DC 20420
- Associate Director, Cohort Management & Public Relations - Jennifer E. Deen, B.S.
  US Department of Veterans Affairs, 810 Vermont Avenue NW, Washington, DC 20420

**MVP Executive Committee**

- Co-Chair: J. Michael Gaziano, M.D., M.P.H.
  VA Boston Healthcare System, 150 S. Huntington Avenue, Boston, MA 02130
- Co-Chair: Sumitra Muralidhar, Ph.D.
  US Department of Veterans Affairs, 810 Vermont Avenue NW, Washington, DC 20420
- Jean Beckham, Ph.D.
  Durham VA Medical Center, 508 Fulton Street, Durham, NC 27705
- Kyong-Mi Chang, M.D.
  Philadelphia VA Medical Center, 3900 Woodland Avenue, Philadelphia, PA 19104
- Philip S. Tsao, Ph.D.
  VA Palo Alto Health Care System, 3801 Miranda Avenue, Palo Alto, CA 94304
- Shiuh-Wen Luoh, M.D., Ph.D.
  VA Portland Health Care System, 3710 SW US Veterans Hospital Rd, Portland, OR 97239
  US Department of Veterans Affairs, 810 Vermont Avenue NW, Washington, DC 20420
- Juan P. Casas, M.D., Ph.D., Ex-Officio
  VA Boston Healthcare System, 150 S. Huntington Avenue, Boston, MA 02130

**MVP Principal Investigators**

- J. Michael Gaziano, M.D., M.P.H.
  VA Boston Healthcare System, 150 S. Huntington Avenue, Boston, MA 02130
- Philip S. Tsao, Ph.D.
  VA Palo Alto Health Care System, 3801 Miranda Avenue, Palo Alto, CA 94304

**MVP Operations**

- MVP Executive Director – Juan P. Casas, M.D., Ph.D.
  VA Boston Healthcare System, 150 S. Huntington Avenue, Boston, MA 02130
- Director of Regulatory Affairs – Lori Churby, B.S.
  VA Palo Alto Health Care System, 3801 Miranda Avenue, Palo Alto, CA 94304
- MVP Cohort Management Director – Stacey B. Whitbourne, Ph.D.
  VA Boston Healthcare System, 150 S. Huntington Avenue, Boston, MA 02130
- MVP Recruitment/Enrollment Director - Jessica V. Brewer, M.P.H.
  VA Boston Healthcare System, 150 S. Huntington Avenue, Boston, MA 02130
- Director, VA Central Biorepository, Boston – Mary T. Brophy M.D., M.P.H.
  VA Boston Healthcare System, 150 S. Huntington Avenue, Boston, MA 02130
- Executive Director for MVP Biorepositories - Luis E. Selva, Ph.D.
  VA Boston Healthcare System, 150 S. Huntington Avenue, Boston, MA 02130
- MVP Informatics, Boston – Shahpoor (Alex) Shayan, M.S.

VA Boston Healthcare System, 150 S. Huntington Avenue, Boston, MA 02130
- Director, MVP Data Operations/Analytics, Boston – Kelly Cho, M.P.H., Ph.D.
  VA Boston Healthcare System, 150 S. Huntington Avenue, Boston, MA 02130
- Director, Center for Computational and Data Science (C-DACS) & Genomics Core – Saiju Pyarajan Ph.D.
  VA Boston Healthcare System, 150 S. Huntington Avenue, Boston, MA 02130
- Director, Molecular Data Core – Philip S. Tsao, Ph.D.
  VA Palo Alto Health Care System, 3801 Miranda Avenue, Palo Alto, CA 94304
- Director, Phenomics Data Core – Kelly Cho, M.P.H, Ph.D.
  VA Boston Healthcare System, 150 S. Huntington Avenue, Boston, MA 02130
- Director, VA Informatics and Computing Infrastructure (VINCI) – Scott L. DuVall, Ph.D.
  VA Salt Lake City Health Care System, 500 Foothill Drive, Salt Lake City, UT 84148
- MVP Coordinating Centers
  - Cooperative Studies Program Clinical Research Pharmacy Coordinating Center, Albuquerque – Todd Connor, Pharm.D.; Dean P. Argyres, B.S., M.S.
    New Mexico VA Health Care System, 1501 San Pedro Drive SE, Albuquerque, NM 87108
  - Genomics Coordinating Center, Palo Alto – Philip S. Tsao, Ph.D.
    VA Palo Alto Health Care System, 3801 Miranda Avenue, Palo Alto, CA 94304
  - MVP Boston Coordinating Center, Boston - J. Michael Gaziano, M.D., M.P.H.
    VA Boston Healthcare System, 150 S. Huntington Avenue, Boston, MA 02130
  - MVP Information Center, Canandaigua – Brady Stephens, M.S.
    Canandaigua VA Medical Center, 400 Fort Hill Avenue, Canandaigua, NY 14424

**Current MVP Local Site Investigators**

- Atlanta VA Medical Center (Peter Wilson, M.D.)
  1670 Clairmont Road, Decatur, GA 30033
- Bay Pines VA Healthcare System (Rachel McArdle, Ph.D.)
  10,000 Bay Pines Blvd Bay Pines, FL 33744
- Birmingham VA Medical Center (Louis Dellitalia, M.D.)
  700 S. 19th Street, Birmingham AL 35233
- Central Western Massachusetts Healthcare System (Kristin Mattocks, Ph.D., M.P.H.)
  421 North Main Street, Leeds, MA 01053
- Cincinnati VA Medical Center (John Harley, M.D., Ph.D.)
  3200 Vine Street, Cincinnati, OH 45220
- Clement J. Zablocki VA Medical Center (Jeffrey Whittle, M.D., M.P.H.)
  5000 West National Avenue, Milwaukee, WI 53295
- VA Northeast Ohio Healthcare System (Frank Jacono, M.D.)
  10701 East Boulevard, Cleveland, OH 44106
- Durham VA Medical Center (Jean Beckham, Ph.D.)
  508 Fulton Street, Durham, NC 27705
- Edith Nourse Rogers Memorial Veterans Hospital (John Wells., Ph.D.)
  200 Springs Road, Bedford, MA 01730
- Edward Hines, Jr. VA Medical Center (Salvador Gutierrez, M.D.)
  5000 South 5th Avenue, Hines, IL 60141
- Veterans Health Care System of the Ozarks (Kathrina Alexander, M.D.)
  1100 North College Avenue, Fayetteville, AR 72703
- Fargo VA Health Care System (Kimberly Hammer, Ph.D.)
  2101 N. Elm, Fargo, ND 58102

- VA Health Care Upstate New York (James Norton, Ph.D.)
  113 Holland Avenue, Albany, NY 12208
- New Mexico VA Health Care System (Gerardo Villareal, M.D.)
  1501 San Pedro Drive, S.E. Albuquerque, NM 87108
- VA Boston Healthcare System (Scott Kinlay, M.B.B.S., Ph.D.)
  150 S. Huntington Avenue, Boston, MA 02130
- VA Western New York Healthcare System (Junzhe Xu, M.D.)
  3495 Bailey Avenue, Buffalo, NY 14215-1199
- Ralph H. Johnson VA Medical Center (Mark Hamner, M.D.)
  109 Bee Street, Mental Health Research, Charleston, SC 29401
- Columbia VA Health Care System (Roy Mathew, M.D.)
  6439 Garners Ferry Road, Columbia, SC 29209
- VA North Texas Health Care System (Sujata Bhushan, M.D.)
  4500 S. Lancaster Road, Dallas, TX 75216
- Hampton VA Medical Center (Pran Iruvanti, D.O., Ph.D.)
  100 Emancipation Drive, Hampton, VA 23667
- Richmond VA Medical Center (Michael Godschalk, M.D.)
  1201 Broad Rock Blvd., Richmond, VA 23249
- Iowa City VA Health Care System (Zuhair Ballas, M.D.)
  601 Highway 6 West, Iowa City, IA 52246-2208
- Eastern Oklahoma VA Health Care System (River Smith, Ph.D.)
  1011 Honor Heights Drive, Muskogee, OK 74401
- James A. Haley Veterans' Hospital (Stephen Mastorides, M.D.)
  13000 Bruce B. Downs Blvd, Tampa, FL 33612
- James H. Quillen VA Medical Center (Jonathan Moorman, M.D., Ph.D.)
  Corner of Lamont & Veterans Way, Mountain Home, TN 37684
- John D. Dingell VA Medical Center (Saib Gappy, M.D.)
  4646 John R Street, Detroit, MI 48201
- Louisville VA Medical Center (Jon Klein, M.D., Ph.D.)
  800 Zorn Avenue, Louisville, KY 40206
- Manchester VA Medical Center (Nora Ratcliffe, M.D.)
  718 Smyth Road, Manchester, NH 03104
- Miami VA Health Care System (Ana Palacio, M.D., M.P.H.)
  1201 NW 16th Street, 11 GRC, Miami FL 33125
- Michael E. DeBakey VA Medical Center (Olaoluwa Okusaga, M.D.)
  2002 Holcombe Blvd, Houston, TX 77030
- Minneapolis VA Health Care System (Maureen Murdoch, M.D., M.P.H.)
  One Veterans Drive, Minneapolis, MN 55417
- N. FL/S. GA Veterans Health System (Peruvemba Sriram, M.D.)
  1601 SW Archer Road, Gainesville, FL 32608
- Northport VA Medical Center (Shing Shing Yeh, Ph.D., M.D.)
  79 Middleville Road, Northport, NY 11768
- Overton Brooks VA Medical Center (Neeraj Tandon, M.D.)
  510 East Stoner Ave, Shreveport, LA 71101
- Philadelphia VA Medical Center (Darshana Jhala, M.D.)
  3900 Woodland Avenue, Philadelphia, PA 19104
- Phoenix VA Health Care System (Samuel Aguayo, M.D.)
  650 E. Indian School Road, Phoenix, AZ 85012
- Portland VA Medical Center (David Cohen, M.D.)
  3710 SW U.S. Veterans Hospital Road, Portland, OR 97239

- Providence VA Medical Center (Satish Sharma, M.D.)
  830 Chalkstone Avenue, Providence, RI 02908
- Richard Roudebush VA Medical Center (Suthat Liangpunsakul, M.D., M.P.H.)
  1481 West 10th Street, Indianapolis, IN 46202
- Salem VA Medical Center (Kris Ann Oursler, M.D.)
  1970 Roanoke Blvd, Salem, VA 24153
- San Francisco VA Health Care System (Mary Whooley, M.D.)
  4150 Clement Street, San Francisco, CA 94121
- South Texas Veterans Health Care System (Sunil Ahuja, M.D.)
  7400 Merton Minter Boulevard, San Antonio, TX 78229
- Southeast Louisiana Veterans Health Care System (Joseph Constans, Ph.D.)
  2400 Canal Street, New Orleans, LA 70119
- Southern Arizona VA Health Care System (Paul Meyer, M.D., Ph.D.)
  3601 S 6th Avenue, Tucson, AZ 85723
- Sioux Falls VA Health Care System (Jennifer Greco, M.D.)
  2501 W 22nd Street, Sioux Falls, SD 57105
- St. Louis VA Health Care System (Michael Rauchman, M.D.)
  915 North Grand Blvd, St. Louis, MO 63106
- Syracuse VA Medical Center (Richard Servatius, Ph.D.)
  800 Irving Avenue, Syracuse, NY 13210
- VA Eastern Kansas Health Care System (Melinda Gaddy, Ph.D.)
  4101 S 4th Street Trafficway, Leavenworth, KS 66048
- VA Greater Los Angeles Health Care System (Agnes Wallbom, M.D., M.S.)
  11301 Wilshire Blvd, Los Angeles, CA 90073
- VA Long Beach Healthcare System (Timothy Morgan, M.D.)
  5901 East 7th Street Long Beach, CA 90822
- VA Maine Healthcare System (Todd Stapley, D.O.)
  1 VA Center, Augusta, ME 04330
- VA New York Harbor Healthcare System (Peter Liang, M.D., M.P.H.)
  423 East 23rd Street, New York, NY 10010
- VA Pacific Islands Health Care System (Daryl Fujii, Ph.D.)
  459 Patterson Rd, Honolulu, HI 96819
- VA Palo Alto Health Care System (Philip Tsao, Ph.D.)
  3801 Miranda Avenue, Palo Alto, CA 94304-1290
- VA Pittsburgh Health Care System (Patrick Strollo, Jr., M.D.)
  University Drive, Pittsburgh, PA 15240
- VA Puget Sound Health Care System (Edward Boyko, M.D.)
  1660 S. Columbian Way, Seattle, WA 98108-1597
- VA Salt Lake City Health Care System (Jessica Walsh, M.D.)
  500 Foothill Drive, Salt Lake City, UT 84148
- VA San Diego Healthcare System (Samir Gupta, M.D., M.S.C.S.)
  3350 La Jolla Village Drive, San Diego, CA 92161
- VA Sierra Nevada Health Care System (Mostaqul Huq, Pharm.D., Ph.D.)
  975 Kirman Avenue, Reno, NV 89502
- VA Southern Nevada Healthcare System (Joseph Fayad, M.D.)
  6900 North Pecos Road, North Las Vegas, NV 89086
- VA Tennessee Valley Healthcare System (Adriana Hung, M.D., M.P.H.)
  1310 24th Avenue, South Nashville, TN 37212
- Washington DC VA Medical Center (Jack Lichy, M.D., Ph.D.)
  50 Irving St, Washington, D. C. 20422

- W.G. (Bill) Hefner VA Medical Center (Robin Hurley, M.D.)
  1601 Brenner Ave, Salisbury, NC 28144
- White River Junction VA Medical Center (Brooks Robey, M.D.)
  163 Veterans Drive, White River Junction, VT 05009
- William S. Middleton Memorial Veterans Hospital (Prakash Balasubramanian, M.D.)
  2500 Overlook Terrace, Madison, WI 53705

### Supplementary Methods

#### Measurement of the instrument, exposure, outcome and other covariates in the Million Veteran Program

Array-based genotyping for MVP participants was completed using the MVP 1.0 custom Axiom array.[1] We used genetic data from the MVP Release 4 dataset, for which methods for genetic quality control have been previously described.[1] For our proposed instrument, we used data on 14 genetic variants across 3 genes (*HMGCR*, *PCSK9* and *LDLR*) for LDL-C and 42 genetic variants for HDL-C (Tables S2-S3). The 14 LDL-C variants were selected from a prior MR study of LDL-C and coronary artery disease conducted in the UK Biobank, where the variants acted as proxies of existing lipid-lowering therapies.[2] For HDL-C, proposed instruments were identified from variants associated with HDL-C at a genome-wide level of significance ($p < 5\times10^{-8}$) in the Global Lipids Genetics Consortium (GLGC). We performed linkage disequilibrium clumping ($R^2 < 0.001$ and clumping window of 10,000 kb) and, to avoid potential exclusion restriction violations, further excluded any variants that were genome-wide significantly associated with LDL-C, apolipoprotein, alcohol intake, body mass index or waist-hip ratio. Detailed information for additional study variables is provided in Table S1.

LDL-C and HDL-C measurements were obtained from casually obtained outpatient blood specimens and identified using records from the Lab Chemistry domain in the VA's corporate data warehouse (CDW) linked to MVP participants.[3,4] LDL-C was estimated using the Freidewald formula.[5] Data on use of a LDL-C lowering therapy were obtained from the Medications domain.[6] The primary outcome was defined as nonfatal or fatal coronary artery disease events (which included myocardial infarction, atherosclerosis, and undergoing angioplasty or revascularization), and was identified using from records in the Outpatient and Inpatient domains in CDW, Centers for Medicare and Medicaid Services (CMS), and the National Death Index[7] using diagnosis codes (ICD-9 and ICD-10; obtained from a prior study conducted in UK Biobank)[2], and procedure codes (ICD-9, ICD-10 and CPT; obtained from a prior study conducted in MVP)[8]. We also conducted sensitivity analyses in which the ICD-9 and ICD-10 diagnosis codes were obtained from a previously study conducted in MVP.[8] Abnormal liver function was measured via alanine transaminase (ALT) or aspartate aminotransferase (ASN) greater than three times the upper limit of normal in the past year.

### **Supplementary References**

1. Hunter-Zinck H, Shi Y, Li M, et al. Genotyping Array Design and Data Quality Control in the Million Veteran Program. *Am J Hum Genet*. 2020 Apr 2;**106**(4):535–548.

2. Allara E, Morani G, Carter P, et al. Genetic Determinants of Lipids and Cardiovascular Disease Outcomes: A Wide-Angled Mendelian Randomization Investigation. *Circ: Genomic and Precision Medicine*. 2019 Dec;**12**(12):e002711.

3. CIPHER - Low Density Lipoprotein- Cholesterol (MVP Data Core) [Internet]. [cited 2024 Aug 21]. Available from: https://phenomics.va.ornl.gov/web/cipher/phenotype-viewer?uqid=69dc3f9bde1947bdb9c54cce615a0f58&name=Low_Density_Lipoprotein-__Cholesterol__MVP_Data_Core_

4. CIPHER - High-Density Lipoprotein Cholesterol (MVP Data Core) [Internet]. [cited 2024 Aug 21]. Available from: https://phenomics.va.ornl.gov/web/cipher/phenotype-viewer?uqid=cd1aa8db704a467492f75a7528d0363e&name=High-Density_Lipoprotein_Cholesterol__MVP_Data_Core_

5. Friedewald WT, Levy RI, Fredrickson DS. Estimation of the concentration of low-density lipoprotein cholesterol in plasma, without use of the preparative ultracentrifuge. *Clin Chem*. 1972 Jun;**18**(6):499–502.

6. CIPHER - Antilipemic Agents, Class (MVP Data Core) [Internet]. [cited 2024 Aug 21]. Available from: https://phenomics.va.ornl.gov/web/cipher/phenotype-viewer?uqid=ab33dcac05954de59e87267fb91b6b05&name=Antilipemic_Agents__Class__MVP_Data_Core_

7. Veterans Health Administration, Center of Excellence for Suicide Prevention. Joint Department of Veteran Affairs and Department of Defense Mortality Data Repository. Data compiled from the National Death Index [Internet]. Available from: https://www.mirecc.va.gov/suicideprevention/documents/VA_DoD-MDF_Flyer.pdf; Extract November 18, 2022

8. Seligman B, Qazi S, Rane M, et al. Association between multiple frailty indices and CVD mortality. *Innovation in Aging*. 2023 Dec 21;**7**(Supplement_1):503–503.

### Supplementary Figures

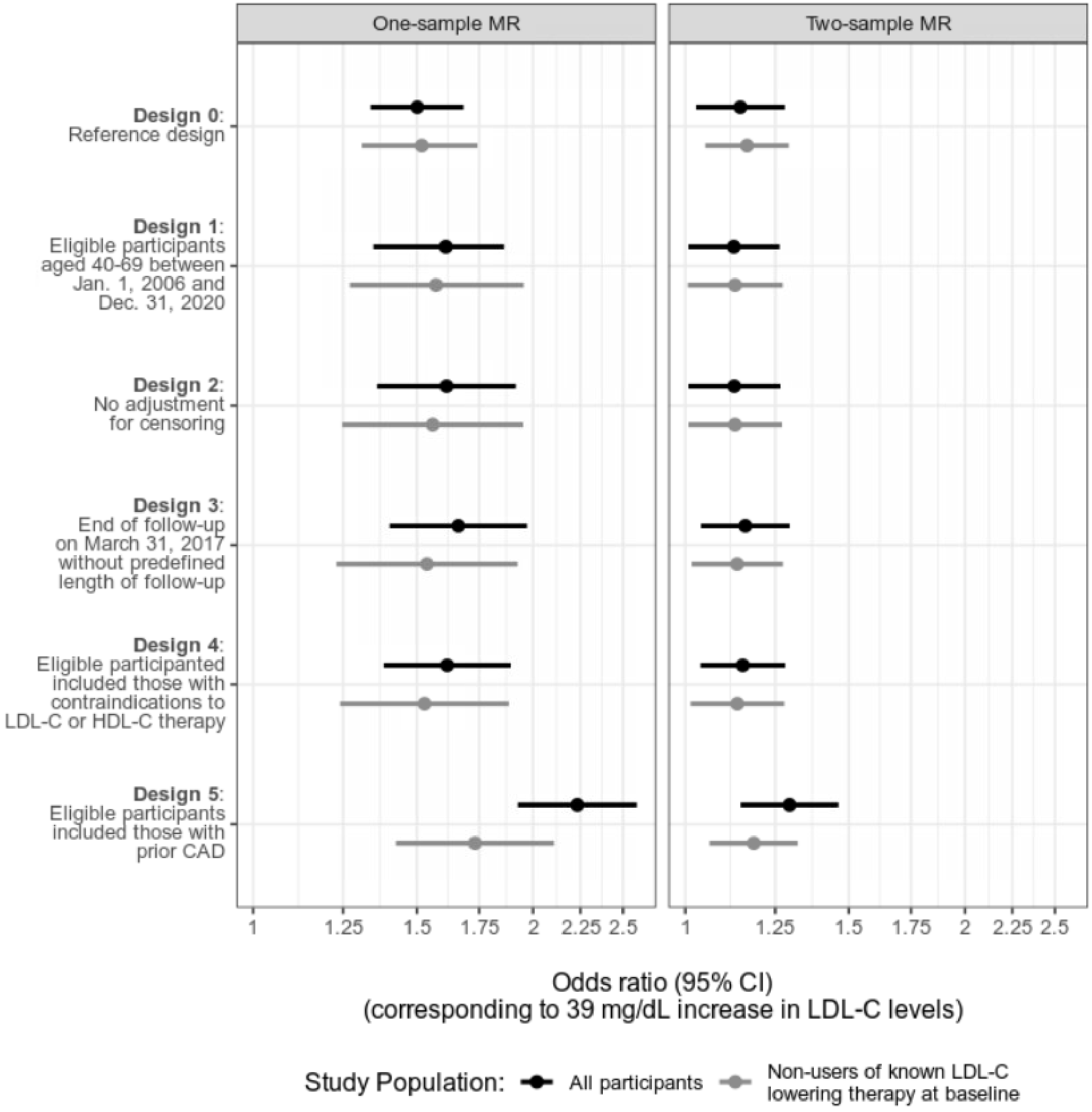

**Figure S1. MR estimates of odds ratios and 95% confidence intervals for LDL-C, by use of LDL-C lowering therapy at the start of follow-up (all participants versus restricted to non-users), Million Veteran Program**

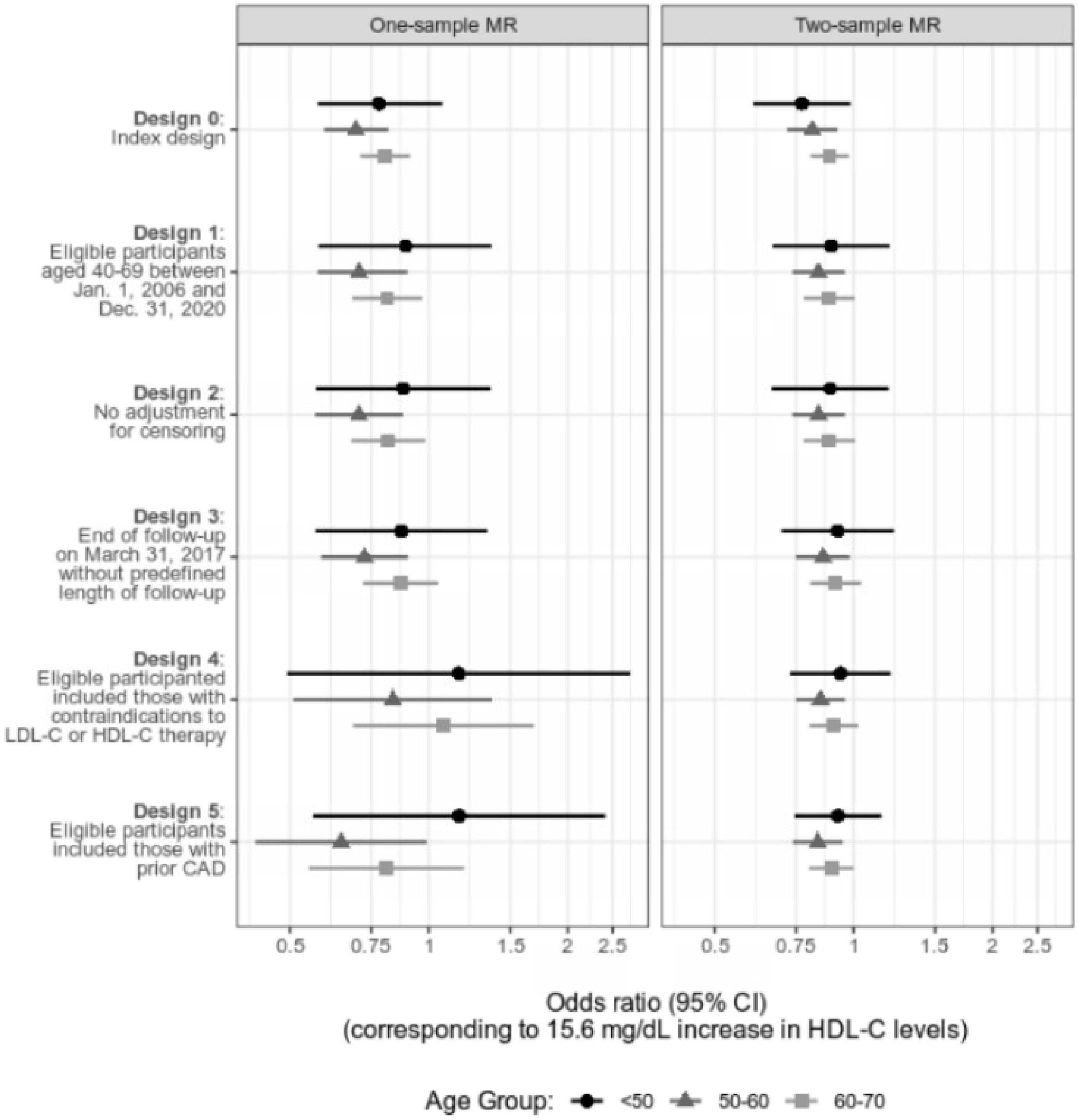

**Figure S2. MR estimates of odds ratios and 95% confidence intervals for HDL-C across different age groups (<50, 50 to <60, 60 to <70), Million Veteran Program**

### Supplementary Tables

**Table S1. Variables used in the Mendelian randomization study of LDL-C, HDL-C and coronary artery disease using observational data from the Million Veteran Program (January 1, 2002 to December 31, 2020)**

| Variable | Variable type | Detail | Codes |
|---|---|---|---|
| Age | Continuous (years) | Based on records in the MVP Roster file. | N/A |
| Sex | Binary:<br>Male / Female | Based on records in the MVP Core Demographics file. | N/A |
| Harmonized ancestry and race/ethnicity (HARE) | Categorical:<br>Non-Hispanic white /<br>Non-Hispanic Black /<br>Non-Hispanic Asian /<br>Hispanic | Genetically defined ancestry based on the use of a supervised machine learning algorithm applied to self-identified racial and ethnic (SIRE) background | N/A |
| Primary care visit in the past year | Binary:<br>Yes / No | Defined as an in-person or telehealth primary care visit, based on stop codes in any position. Based on records in the *Outpatient* domain. | **Stop codes**: 156, 157, 170-178, 322, 323, 338, 341, 342, 348, 350, 531, 534, 539, 704 |
| Hospitalization in the past 12 months | Binary:<br>Yes / No | Defined as receipt of inpatient care. Based on records in the *Inpatient* domain. | N/A |
| Smoking status | Categorical:<br>Never / Current<br>Former | Defined as the most recent self-reported smoking status prior to the start of follow-up. Based on records in the *Health Factors* domain. | N/A |
| Body mass index | Continuous ($kg/m^2$) | Calculated using the most recent height and weight measurements prior to the start of follow-up. Based on records in the *Vital* domain. | N/A |
| Systolic blood pressure | Continuous (mmHg) | Defined as the most recent measurement prior to the start of follow-up. Based on records in the *Vital* domain. | N/A |
| Diastolic blood pressure | Continuous (mmHg) | Defined as the most recent measurement prior to the start of follow-up. Based on records in the *Vital* domain. | N/A |
| Total cholesterol | Continuous (mg/dL) | Defined as the most recent measurement of total cholesterol prior to the start of follow-up, identified using an adjudicated list. Based on records in the *Labs* domain. | **Short Name**: TotChol |

| Variable | Variable type | Detail | Codes |
|---|---|---|---|
| Triglycerides | Continuous (mg/dL) | Defined as the most recent measurement of triglycerides prior to the start of follow-up, identified using an adjudicated list. Based on records in the *Labs* domain. | **Short Name**: Trig |
| High-density lipoprotein cholesterol (HDL-C) | Continuous (mg/dL) | Defined as the most recent measurement of HDL-C prior to the start of follow-up, identified using an adjudicated list. Based on records in the *Labs* domain. | **Short Name**: HDLC |
| Low-density lipoprotein cholesterol (LDL-C) | Continuous (mg/dL) | Defined as the most recent measurement of LDL-C prior to the start of follow-up, identified using an adjudicated list. Based on records in the *Labs* domain. | **Short Name**: LDLC |
| Alanine transaminase (ALT) | Continuous (IU/L) | Defined as the most recent measurement of ALT prior to the start of follow-up, identified using an adjudicated list. Based on records in the *Labs* domain. | **Short Name**: ALT |
| Aspartate aminotransferase (AST) | Continuous (IU/L) | Defined as the most recent measurement of AST prior to the start of follow-up, identified using an adjudicated list. Based on records in the *Labs* domain. | **Short Name**: AST |
| Use of a LDL-C lowering therapy in the past year | Binary: Yes / No | LDL-C lowering therapies, identified using an adjudicated list based on the following generic names: colestipol, dextrophyroxine, evolocumab, ezetimibe, ezetimibe/simvastatin, fenofibrate, fenofibric acid, fish oil, gemfibrozil, icosapent ethyl, lomitapide, mevacor, mipomersen niacinamide, omega-3, omega-3 acid, probucol, rosuvastatin, statin, alirocumab, atorvastatin, atromid, bezafibrate, bezalip retard, cholestyramine, choloxin, clofibrate, colesevelam. Based on records in the *Medications* domain. | **VA Classification**: CV350 |
| Coronary artery disease (primary definition) | Binary: Yes / No | Defined as ≥1 diagnosis or procedure code in the *Inpatient* domain or ≥2 diagnosis codes in the *Outpatient* domain | **ICD-9**: 41[0-2].%, 414.0%, 414.[8-9]%<br>**ICD-10**: I2[1-4].%, I25.1%, 125.2, 125.5, 125.6, 125.8%, I25.9 |

| Variable | Variable type | Detail | Codes |
|---|---|---|---|
| | | | **ICD-9 Procedure**: 36.01, 36.02, 36.05, 36.1%, 00.66<br>**ICD-10 Procedure**: 021008W, 021[0-1]09[CFW], 021[0-2]0A[389CW], 02100J[389W], 0200K[389W], 021[0-1]0Z%, 021[0-2]49W, 02104A[89W], 02104J3, 02104K[9W], 02104Z[389], 02110J[9W], 02110KW, 02114AW, 02114J9, 02114K3, 02114Z[39], 021209[CW], 021[2-3]0J[389W], 02120K[39W], 02120Z[389C], 02124A9, 02124Z3, 021309W, 02130A[89W], 02130KW, 02130Z[39], 021349C, 02134KW, 02134Z3, 02703[4567DEFGTZ]Z, 02703[DZ]6, 02704[4567DEFGZ]Z, 02704[DZ]6, 02713[4567DEFGZ]Z, 02713[DFGZ]6, 02714[4567DEGZ]Z, 02723[4567DEFGZ]Z, 02723[DFGZ]6, 02724[4567DEFZ]Z, 02733[4567DGZ]Z, 02733[GZ]6, 02734[4567DGZ]Z<br>**CPT**: 33150-33536, 9292% 9293% 9294% , 92974, 92975 |
| Coronary artery disease (secondary definition) | Binary:<br>Yes / No | Defined as ≥1 diagnosis or procedure code in the *Inpatient* domain or ≥2 diagnosis codes in the *Outpatient* domain | **ICD-9**: 41[0-2].%, 414.%, 429.2, 429.5, 429.7%, V45.82<br>**ICD-10**: I20.0, I2[1-5].% (excluding I25.112 and I25.7[0-9]2), I51.[0-1]%, Z95.5%, Z98.61<br>**ICD-9 Procedure**: 36.01, 36.02, 36.05, 36.1%, 00.66<br>**ICD-10 Procedure**: 021008W, 021[0-1]09[CFW], 021[0-2]0A[389CW], 02100J[389W], 0200K[389W], 021[0-1]0Z%, 021[0-2]49W, 02104A[89W], 02104J3, 02104K[9W], 02104Z[389], 02110J[9W], 02110KW, 02114AW, 02114J9, 02114K3, 02114Z[39], 021209[CW], 021[2-3]0J[389W], 02120K[39W], 02120Z[389C], 02124A9, 02124Z3, |

| Variable | Variable type | Detail | Codes |
|---|---|---|---|
| | | | 021309W, 02130A[89W], 02130KW, 02130Z[39], 021349C, 02134KW, 02134Z3, 02703[4567DEFGTZ]Z, 02703[DZ]6, 02704[4567DEFGZ]Z, 02704[DZ]6, 02713[4567DEFGZ]Z, 02713[DFGZ]6, 02714[4567DEGZ]Z, 02723[4567DEFGZ]Z, 02723[DFGZ]6, 02724[4567DEFZ]Z, 02733[4567DGZ]Z, 02733[GZ]6, 02734[4567DGZ]Z<br>**CPT**: 33150-33536, 929[2-4]% , 92974, 92975 |
| Myopathy | Binary:<br>Yes / No | Defined as ≥1 diagnosis in the *Inpatient* domain or ≥2 diagnoses in the *Outpatient* domain | **ICD-9**: 359.[4-9]%<br>**ICD-10**: G71.3, G71.[8-9]%, G72.[0-2]%, G72.4%, G72.[8-9]%, G73.7, M05.4[0-7]%, M05.49, M33.[0-2]2, M33.92, M34.82, M35.03, M60.%, M62.9 |
| Chronic liver disease | Binary:<br>Yes / No | Defined as ≥1 diagnosis in the *Inpatient* domain or ≥2 diagnoses in the *Outpatient* domain | **ICD-9**: 571.%, 572.%, 573.3%, 070.%<br>**ICD-10**: K70.%, K7[2-5].% |
| Chronic kidney disease or end renal disease | Binary:<br>Yes / No | Defined as ≥1 diagnosis in the *Inpatient* domain or ≥2 diagnoses in the *Outpatient* domain | **ICD-9**: 016.0%, 095.4, 189.0, 189.9, 223.0, 236.91, 249.4%, 250.4%, 271.4, 274.10, 283.11, 403.[0-1]%, 403.91%, 404.0[2-3]%, 404.1[2-3]%, 404.9[2-3]%, 440.1, 442.1, 572.4, 580[0-8].%, 581.%, 591.%, 573.1[2-9]%, 573.2%, 794.4<br>**ICD-10**: A18.11, A52.75, B52.0, C64.%, C68.9, D30.0%, D41.[0-2]%, D59.3, E08.2%, E08.65, E09.2%, E10.2%, E10.65, E11.2%, E11.65, E13.2%, E74.8, I1[2-3].%, I70.1, I72.2, K76.7, M10.3%, M32.1[4-5], M35.04, N0.%, N13.[1-3]%, N14.%, N15.0, N15.[8-9], N1[6-9], N25.%, N26.1, N26.9, Q61.02, Q61.[1-5]%, Q61.8, Q62.[0-3]%, R94.4<br>**ICD-9 Procedure**: 38.95, 39.93, 39.95, 54.98%, 99.78 |

| Variable | Variable type | Detail | Codes |
|---|---|---|---|
| | | | **ICD-10 Procedure**: 031[3-8]0JD, 03190JF, 031[ABC]0JF, 3E1M39Z, 5A1D[7-9]0Z, 6ABT0BZ |
| Loss to follow-up | Binary:<br>Yes / No | Defined as one year after the last recorded LDL-C or HDL-C measurement or one year after the last visit to a primary care physician, whichever occurred later | N/A |

**Table S2. Summary statistics for the associations between the proposed instruments for LDL-C (based on genetic variants in the *HMGCR*, *PCSK9* and *LDLR* genes) and LDL-C, obtained from the Global Lipids Genetics Consortium**

| Gene | SNP | Position | Reference allele | Alternate allele | Beta | SE | Reference allele frequency |
|---|---|---|---|---|---|---|---|
| *PCSK9* | rs2479394 | chr1:55258652 | G | A | 0.0386 | 0.0041 | 0.2850 |
| *PCSK9* | rs11206510 | chr1:55268627 | T | C | 0.0831 | 0.0050 | 0.8456 |
| *PCSK9* | rs2149041 | chr1:55274725 | G | C | 0.0636 | 0.0049 | 0.1609 |
| *PCSK9* | rs10888897 | chr1:55285649 | C | T | 0.0507 | 0.0042 | 0.6055 |
| *PCSK9* | rs7552841 | chr1:55291340 | T | C | 0.0368 | 0.0044 | 0.3654 |
| *PCSK9* | rs562556 | chr1:55296825 | A | G | 0.0640 | 0.0066 | 0.8061 |
| *HMGCR* | rs10066707 | chr5:74596335 | A | G | 0.0497 | 0.0054 | 0.4169 |
| *HMGCR* | rs2006760 | chr5:74597785 | G | C | 0.0533 | 0.0076 | 1.0000 |
| *HMGCR* | rs2303152 | chr5:74677463 | A | G | 0.0423 | 0.0064 | 0.1201 |
| *HMGCR* | rs17238484 | chr5:74684252 | T | G | 0.0627 | 0.0062 | 0.2533 |
| *HMGCR* | rs5909 | chr5:74691931 | A | G | 0.0617 | 0.0088 | 0.1016 |
| *HMGCR* | rs12916 | chr5:74692295 | C | T | 0.0733 | 0.0038 | 0.4314 |
| *LDLR* | rs6511720 | chr19:11063306 | G | T | 0.2209 | 0.0061 | 0.9024 |
| *LDLR* | rs688 | chr19:11088602 | T | C | 0.0540 | 0.0037 | 0.4472 |

**Table S3. Summary statistics for the associations between the proposed instruments for HDL-C and HDL-C, obtained from the Global Lipids Genetics Consortium**

| SNP | Position | Reference allele | Alternate allele | Beta | SE | Reference allele frequency |
|---|---|---|---|---|---|---|
| rs12145743 | chr1:156700651 | G | T | 0.0203 | 0.0036 | 0.3311 |
| rs4650994 | chr1:178515312 | G | A | 0.0210 | 0.0034 | 0.5172 |
| rs1689797 | chr1:182150978 | C | A | 0.0358 | 0.0036 | 0.6979 |
| rs4660293 | chr1:40028180 | A | G | 0.0353 | 0.0040 | 0.7639 |
| rs12133576 | chr1:93816400 | A | G | 0.0243 | 0.0035 | 0.3549 |
| rs1515110 | chr2:227122216 | G | T | 0.0323 | 0.0035 | 0.3813 |
| rs6805251 | chr3:119560606 | T | C | 0.0200 | 0.0035 | 0.3813 |
| rs13076253 | chr3:131751775 | A | C | 0.0283 | 0.0048 | 0.8522 |
| rs13099479 | chr3:52677478 | A | G | 0.0360 | 0.0062 | 0.0897 |
| rs2602836 | chr4:100014805 | A | G | 0.0192 | 0.0034 | 0.4274 |
| rs3822072 | chr4:89741269 | G | A | 0.0251 | 0.0034 | 0.5119 |
| rs6450176 | chr5:53298025 | G | A | 0.0254 | 0.0039 | 0.7216 |
| rs3861397 | chr6:139828916 | A | G | 0.0240 | 0.0036 | 0.6583 |
| rs9457931 | chr6:160929904 | A | G | 0.0552 | 0.0073 | 0.9314 |
| rs1980493 | chr6:32363215 | T | C | 0.0318 | 0.0048 | 0.8773 |
| rs11765979 | chr7:130445877 | C | A | 0.0412 | 0.0048 | 0.4578 |
| rs17173637 | chr7:150529449 | T | C | 0.0363 | 0.0057 | 0.9024 |
| rs10087900 | chr8:144303418 | G | A | 0.0231 | 0.0036 | 0.5607 |
| rs12412743 | chr10:114045333 | C | T | 0.0291 | 0.0045 | 0.847 |
| rs970548 | chr10:46013277 | C | A | 0.0258 | 0.0039 | 0.277 |
| rs10761771 | chr10:65230164 | C | T | 0.0198 | 0.0034 | 0.467 |
| rs7112577 | chr11:117044603 | G | C | 0.0826 | 0.0129 | 1.000 |
| rs10501321 | chr11:47294626 | C | T | 0.0483 | 0.0036 | 0.314 |
| rs12801636 | chr11:65391317 | A | G | 0.0235 | 0.0042 | 0.2243 |
| rs2241210 | chr12:109950144 | G | A | 0.0332 | 0.0035 | 0.5528 |
| rs2454722 | chr12:123171218 | G | A | 0.0351 | 0.0044 | 0.1451 |
| rs838876 | chr12:125259888 | A | G | 0.0493 | 0.0039 | 0.3259 |
| rs7306660 | chr12:125327384 | G | A | 0.0345 | 0.0036 | 0.6306 |
| rs4379922 | chr12:125351116 | C | T | 0.0247 | 0.0036 | 0.3496 |
| rs11045163 | chr12:20463526 | G | A | 0.0217 | 0.0035 | 0.4063 |
| rs4983559 | chr14:105277209 | G | A | 0.0197 | 0.0036 | 0.3773 |
| rs492571 | chr15:44211273 | T | C | 0.0663 | 0.0090 | 0.9578 |
| rs424346 | chr15:59010962 | T | C | 0.0679 | 0.0113 | 0.0488 |
| rs2241770 | chr16:56866196 | T | C | 0.0989 | 0.0057 | 0.8971 |
| rs16965220 | chr16:57065121 | A | C | 0.0219 | 0.0037 | 0.2982 |
| rs4148005 | chr17:66882466 | T | G | 0.0283 | 0.0036 | 0.7005 |
| rs4969178 | chr17:76388202 | G | A | 0.0263 | 0.0035 | 0.6266 |
| rs2288912 | chr19:45449199 | G | C | 0.0297 | 0.0036 | 0.4987 |
| rs2278236 | chr19:8431581 | A | G | 0.0331 | 0.0035 | 0.5435 |
| rs6031587 | chr20:43038249 | C | T | 0.0488 | 0.0074 | 0.9314 |
| rs4465830 | chr20:44585420 | A | G | 0.0597 | 0.0044 | 0.7982 |
| rs181360 | chr22:21928916 | T | G | 0.0376 | 0.0042 | 0.8008 |

**Table S4. Number of participants and coronary artery disease events for each MR design, stratified by age group (<50, 50 to <60, 60 to <70) with odds ratios and 95% confidence intervals corresponding to a 39 mg/dL increase in LDL-C obtained via one-sample and two-sample approaches**

| MR Design: Modifications to reference design | Age subgroup | Number of participants | Number of events | Odds ratio (95% CI) corresponding to a 39 mg/dL increase in LDL-C | | | |
|---|---|---|---|---|---|---|---|
| | | | | One-sample | Two-sample | | |
| | | | | Two-stage least squares | Inverse-variance weighting | MR-Egger regression | Weighted median regression |
| **Design 0**: Reference MR design | All | 365,725 | 68,088 | 1.50 (1.34, 1.68) | 1.15 (1.03, 1.28) | 1.35 (1.10, 1.66) | 1.27 (1.16, 1.39) |
| | ≤50 | 110,576 | 7,300 | 1.39 (1.11, 1.73) | 1.07 (0.88, 1.29) | 1.21 (0.81, 1.80) | 1.06 (0.86, 1.31) |
| | >50 to ≤60 | 162,986 | 28,676 | 1.51 (1.29, 1.79) | 1.18 (1.07, 1.31) | 1.36 (1.12, 1.66) | 1.22 (1.09, 1.37) |
| | >60 to ≤70 | 197,496 | 45,528 | 1.59 (1.33, 1.95) | 1.12 (1.00, 1.26) | 1.34 (1.07, 1.66) | 1.16 (1.04, 1.30) |
| **Design 1**: Eligible participants aged 40-69 between Jan. 1, 2006 and Dec. 31, 2010 | All | 179,449 | 37,739 | 1.61 (1.35, 1.86) | 1.13 (1.01, 1.26) | 1.36 (1.11, 1.67) | 1.22 (1.10, 1.35) |
| | ≤50 | 36,104 | 3,873 | 1.20 (0.83, 1.71) | 1.04 (0.85, 1.28) | 1.05 (0.69, 1.59) | 1.09 (0.83, 1.44) |
| | >50 to ≤60 | 85,673 | 17,298 | 1.76 (1.38, 2.23) | 1.21 (1.07, 1.38) | 1.44 (1.12, 1.84) | 1.33 (1.16, 1.53) |
| | >60 to ≤70 | 99,634 | 25,800 | 1.60 (1.20, 2.07) | 1.10 (0.98, 1.23) | 1.38 (1.13, 1.66) | 1.22 (1.07, 1.39) |
| **Design 2**: No adjustment for censoring (due to loss to or administrative end of follow-up) | All | 179,449 | 37,739 | 1.61 (1.36, 1.92) | 1.13 (1.01, 1.27) | 1.36 (1.10, 1.67) | 1.22 (1.10, 1.35) |
| | ≤50 | 36,104 | 3,873 | 1.18 (0.82, 1.71) | 1.04 (0.85, 1.28) | 1.04 (0.68, 1.58) | 1.09 (0.83, 1.44) |
| | >50 to ≤60 | 85,673 | 17,298 | 1.76 (1.38, 2.25) | 1.21 (1.07, 1.38) | 1.44 (1.12, 1.84) | 1.33 (1.15, 1.53) |
| | >60 to ≤70 | 99,634 | 25,800 | 1.58 (1.21, 2.07) | 1.10 (0.97, 1.23) | 1.37 (1.12, 1.68) | 1.23 (1.08, 1.40) |
| **Design 3**: End of follow-up on March 31, 2017 without predefined length of follow-up | All | 179,449 | 38,892 | 1.66 (1.40, 1.97) | 1.16 (1.04, 1.30) | 1.38 (1.12, 1.69) | 1.26 (1.14, 1.40) |
| | ≤50 | 36,104 | 3,939 | 1.09 (0.76, 1.56) | 1.05 (0.85, 1.29) | 0.98 (0.64, 1.49) | 1.05 (0.80, 1.38) |
| | >50 to ≤60 | 85,673 | 18,029 | 1.81 (1.43, 2.30) | 1.24 (1.10, 1.39) | 1.40 (1.11, 1.78) | 1.31 (1.14, 1.51) |
| | >60 to ≤70 | 99,634 | 24,799 | 1.67 (1.27, 2.20) | 1.12 (1.01, 1.25) | 1.38 (1.15, 1.65) | 1.25 (1.10, 1.43) |
| **Design 4**: Eligible participants included those with contraindication to LDL-C or HDL-C therapy | All | 194,035 | 42,602 | 1.62 (1.38, 1.89) | 1.15 (1.04, 1.28) | 1.36 (1.12, 1.65) | 1.28 (1.16, 1.42) |
| | ≤50 | 39,354 | 4,452 | 1.17 (0.84, 1.63) | 1.05 (0.86, 1.27) | 1.01 (0.67, 1.52) | 1.13 (0.87, 1.46) |
| | >50 to ≤60 | 95,742 | 20,385 | 1.71 (1.38, 2.12) | 1.23 (1.10, 1.37) | 1.37 (1.09, 1.71) | 1.29 (1.12, 1.47) |
| | >60 to ≤70 | 107,480 | 27,054 | 1.60 (1.23, 2.07) | 1.11 (1.00, 1.22) | 1.35 (1.13, 1.60) | 1.24 (1.10, 1.41) |
| **Design 5**: Eligible participants included those with prior coronary artery disease | All | 235,829 | 84,396 | 2.23 (1.93, 2.59) | 1.30 (1.15, 1.46) | 1.58 (1.26, 1.97) | 1.46 (1.34, 1.59) |
| | ≤50 | 41,937 | 7,035 | 1.41 (1.06, 1.89) | 1.19 (1.00, 1.43) | 1.23 (0.84, 1.79) | 1.23 (0.99, 1.52) |
| | >50 to ≤60 | 114,418 | 39,061 | 2.27 (1.86, 2.76) | 1.36 (1.19, 1.55) | 1.66 (1.29, 2.13) | 1.46 (1.30, 1.63) |
| | >60 to ≤70 | 142,031 | 61,605 | 2.44 (1.91, 3.12) | 1.25 (1.10, 1.41) | 1.56 (1.25, 1.94) | 1.46 (1.31, 1.62) |

**Table S5. Number of participants and coronary artery disease events after restricting to participants with no prior use of a known LDL-C lowering therapy in the 12 months before the start of follow-up for each MR design, stratified by age group (<50, 50 to <60, 60 to <70) with odds ratios and 95% confidence intervals corresponding to a 39 mg/dL increase in LDL-C obtained via one-sample and two-sample approaches**

| MR Design: Modifications to reference design | Age subgroup | Number of participants | Number of events | Odds ratio (95% CI) corresponding to a 39 mg/dL increase in LDL-C | | | |
|---|---|---|---|---|---|---|---|
| | | | | One-sample | Two-sample | | |
| | | | | Two-stage least squares | Inverse-variance weighting | MR-Egger regression | Weighted median regression |
| **Design 0**: Reference MR design | All | 276,101 | 42,247 | 1.52 (1.31, 1.74) | 1.17 (1.05, 1.29) | 1.38 (1.14, 1.66) | 1.27 (1.13, 1.41) |
| | ≤50 | 96,696 | 5,002 | 1.24 (0.96, 1.65) | 1.03 (0.86, 1.23) | 1.17 (0.82, 1.69) | 1.03 (0.81, 1.31) |
| | >50 to ≤60 | 110,456 | 16,340 | 1.38 (1.11, 1.70) | 1.20 (1.06, 1.35) | 1.31 (1.02, 1.67) | 1.20 (1.04, 1.39) |
| | >60 to ≤70 | 117,959 | 24,013 | 1.60 (1.26, 2.01) | 1.14 (1.02, 1.28) | 1.30 (1.04, 1.62) | 1.23 (1.08, 1.40) |
| **Design 1**: Eligible participants aged 40-69 between Jan. 1, 2006 and Dec. 31, 2010 | All | 106,524 | 18,941 | 1.57 (1.27, 1.96) | 1.13 (1.01, 1.27) | 1.31 (1.05, 1.65) | 1.26 (1.10, 1.44) |
| | ≤50 | 25,662 | 2,245 | 1.01 (0.63, 1.61) | 0.90 (0.69, 1.17) | 1.12 (0.66, 1.92) | 0.97 (0.69, 1.38) |
| | >50 to ≤60 | 48,392 | 8,236 | 1.28 (0.92, 1.78) | 1.15 (0.97, 1.37) | 1.30 (0.92, 1.85) | 1.17 (0.96, 1.42) |
| | >60 to ≤70 | 49,899 | 11,519 | 1.79 (1.21, 2.43) | 1.14 (0.98, 1.31) | 1.31 (0.98, 1.75) | 1.27 (1.06, 1.52) |
| **Design 2**: No adjustment for censoring (due to loss to or administrative end of follow-up) | All | 106,524 | 18,941 | 1.56 (1.25, 1.95) | 1.13 (1.01, 1.27) | 1.30 (1.04, 1.63) | 1.25 (1.08, 1.43) |
| | ≤50 | 25,662 | 2,245 | 0.99 (0.61, 1.62) | 0.89 (0.68, 1.17) | 1.10 (0.65, 1.89) | 0.96 (0.68, 1.37) |
| | >50 to ≤60 | 48,392 | 8,236 | 1.28 (0.94, 1.75) | 1.16 (0.97, 1.37) | 1.30 (0.92, 1.85) | 1.17 (0.96, 1.42) |
| | >60 to ≤70 | 49,899 | 11,519 | 1.76 (1.22, 2.54) | 1.13 (0.98, 1.30) | 1.29 (0.97, 1.72) | 1.25 (1.05, 1.49) |
| **Design 3**: End of follow-up on March 31, 2017 without predefined length of follow-up | All | 106,524 | 18,782 | 1.54 (1.23, 1.93) | 1.14 (1.02, 1.27) | 1.23 (0.98, 1.56) | 1.20 (1.05, 1.38) |
| | ≤50 | 25,662 | 2,231 | 0.94 (0.58, 1.55) | 0.88 (0.67, 1.15) | 1.06 (0.62, 1.82) | 0.97 (0.68, 1.37) |
| | >50 to ≤60 | 48,392 | 8,357 | 1.27 (0.93, 1.73) | 1.12 (0.96, 1.29) | 1.18 (0.88, 1.58) | 1.09 (0.90, 1.33) |
| | >60 to ≤70 | 49,899 | 10,738 | 1.82 (1.25, 2.64) | 1.18 (1.03, 1.36) | 1.25 (0.94, 1.67) | 1.30 (1.09, 1.55) |
| **Design 4**: Eligible participants included those with contraindication to LDL-C or HDL-C therapy | All | 116,548 | 20,851 | 1.53 (1.24, 1.88) | 1.14 (1.01, 1.28) | 1.24 (0.98, 1.57) | 1.20 (1.06, 1.37) |
| | ≤50 | 28,146 | 2,548 | 1.03 (0.65, 1.63) | 0.89 (0.69, 1.14) | 1.12 (0.67, 1.85) | 1.07 (0.77, 1.48) |
| | >50 to ≤60 | 55,289 | 9,701 | 1.28 (0.97, 1.70) | 1.13 (0.98, 1.30) | 1.22 (0.91, 1.64) | 1.13 (0.94, 1.36) |
| | >60 to ≤70 | 53,826 | 11,689 | 1.74 (1.22, 2.48) | 1.17 (1.03, 1.33) | 1.20 (0.91, 1.59) | 1.28 (1.08, 1.52) |
| **Design 5**: Eligible participants included those with prior coronary artery disease | All | 124,987 | 29,290 | 1.73 (1.42, 2.11) | 1.19 (1.06, 1.32) | 1.34 (1.08, 1.66) | 1.28 (1.14, 1.43) |
| | ≤50 | 28,817 | 3,219 | 0.97 (0.64, 1.47) | 0.90 (0.72, 1.13) | 0.98 (0.62, 1.54) | 0.94 (0.70, 1.27) |
| | >50 to ≤60 | 58,506 | 12,918 | 1.55 (1.19, 2.02) | 1.19 (1.06, 1.35) | 1.34 (1.05, 1.72) | 1.23 (1.04, 1.44) |
| | >60 to ≤70 | 59,777 | 17,640 | 2.11 (1.50, 2.96) | 1.21 (1.06, 1.38) | 1.39 (1.07, 1.80) | 1.32 (1.14, 1.53) |

**Table S6. Number of participants and coronary artery disease events for each MR design, stratified by age group (<50, 50 to <60, 60 to <70) with odds ratios and 95% confidence intervals corresponding to a 15.6 mg/dL increase in HDL-C obtained via one-sample and two-sample approaches**

| MR Design: Modifications to reference design | Age subgroup | Number of participants | Number of events | Odds ratio (95% CI) corresponding to a 15.6 mg/dL increase in HDL-C | | | |
|---|---|---|---|---|---|---|---|
| | | | | One-sample | Two-sample | | |
| | | | | Two-stage least squares | Inverse-variance weighting | MR-Egger regression | Weighted median regression |
| **Design 0**: Reference MR design | All | 366,515 | 68,319 | 0.76 (0.68, 0.86) | 0.85 (0.76, 0.94) | 0.86 (0.66, 1.12) | 0.87 (0.78, 0.97) |
| | ≤50 | 111,174 | 7,381 | 0.78 (0.57, 1.07) | 0.77 (0.61, 0.98) | 0.72 (0.39, 1.32) | 0.79 (0.60, 1.04) |
| | >50 to ≤60 | 163,897 | 28,912 | 0.69 (0.59, 0.82) | 0.81 (0.72, 0.92) | 0.89 (0.65, 1.21) | 0.91 (0.79, 1.06) |
| | >60 to ≤70 | 197,837 | 45,658 | 0.80 (0.71, 0.91) | 0.89 (0.80, 0.98) | 0.92 (0.72, 1.17) | 0.88 (0.78, 1.00) |
| **Design 1**: Eligible participants aged 40-69 between Jan. 1, 2006 and Dec. 31, 2010 | All | 179,688 | 37,794 | 0.80 (0.69, 0.92) | 0.87 (0.78, 0.96) | 0.98 (0.77, 1.26) | 0.85 (0.75, 0.96) |
| | ≤50 | 36,211 | 3,883 | 0.89 (0.58, 1.37) | 0.89 (0.67, 1.20) | 1.07 (0.52, 2.22) | 1.00 (0.69, 1.45) |
| | >50 to ≤60 | 85,944 | 17,370 | 0.71 (0.57, 0.90) | 0.84 (0.74, 0.96) | 1.09 (0.79, 1.49) | 0.88 (0.74, 1.05) |
| | >60 to ≤70 | 99,711 | 25,819 | 0.81 (0.68, 0.97) | 0.89 (0.78, 1.00) | 0.91 (0.66, 1.24) | 0.95 (0.81, 1.11) |
| **Design 2**: No adjustment for censoring (due to loss to or administrative end of follow-up) | All | 179,688 | 37,794 | 0.79 (0.68, 0.92) | 0.87 (0.78, 0.96) | 0.98 (0.77, 1.26) | 0.85 (0.75, 0.97) |
| | ≤50 | 36,211 | 3,883 | 0.88 (0.57, 1.36) | 0.89 (0.66, 1.19) | 1.07 (0.52, 2.22) | 1.00 (0.69, 1.45) |
| | >50 to ≤60 | 85,944 | 17,370 | 0.71 (0.57, 0.88) | 0.84 (0.74, 0.96) | 1.09 (0.80, 1.50) | 0.86 (0.72, 1.02) |
| | >60 to ≤70 | 99,711 | 25,819 | 0.82 (0.68, 0.98) | 0.89 (0.78, 1.01) | 0.91 (0.66, 1.25) | 0.93 (0.80, 1.09) |
| **Design 3**: End of follow-up on March 31, 2017 without predefined length of follow-up | All | 179,688 | 38,985 | 0.81 (0.70, 0.94) | 0.88 (0.79, 0.98) | 1.05 (0.81, 1.38) | 0.93 (0.82, 1.04) |
| | ≤50 | 36,211 | 3,954 | 0.87 (0.57, 1.34) | 0.92 (0.70, 1.22) | 1.12 (0.55, 2.24) | 1.06 (0.75, 1.51) |
| | >50 to ≤60 | 85,944 | 18,111 | 0.73 (0.58, 0.90) | 0.86 (0.75, 0.98) | 1.14 (0.83, 1.58) | 0.89 (0.74, 1.06) |
| | >60 to ≤70 | 99,711 | 24,836 | 0.87 (0.72, 1.05) | 0.91 (0.80, 1.04) | 1.02 (0.74, 1.40) | 0.95 (0.81, 1.11) |
| **Design 4**: Eligible participants included those with contraindication to LDL-C or HDL-C therapy | All | 194,265 | 42,697 | 0.93 (0.65, 1.34) | 0.87 (0.79, 0.97) | 0.99 (0.77, 1.28) | 0.92 (0.82, 1.03) |
| | ≤50 | 39,470 | 4,468 | 1.16 (0.49, 2.73) | 0.94 (0.73, 1.20) | 0.99 (0.53, 1.85) | 0.98 (0.70, 1.37) |
| | >50 to ≤60 | 96,025 | 20,473 | 0.84 (0.51, 1.37) | 0.85 (0.75, 0.96) | 1.04 (0.77, 1.40) | 0.88 (0.75, 1.04) |
| | >60 to ≤70 | 107,556 | 27,092 | 1.08 (0.68, 1.69) | 0.91 (0.80, 1.02) | 1.01 (0.74, 1.37) | 0.91 (0.79, 1.07) |
| **Design 5**: Eligible participants included those | All | 236,020 | 84,452 | 0.78 (0.57, 1.07) | 0.87 (0.78, 0.97) | 1.02 (0.78, 1.34) | 0.93 (0.84, 1.03) |
| | ≤50 | 42,059 | 7,057 | 1.16 (0.56, 2.41) | 0.92 (0.75, 1.15) | 1.01 (0.59, 1.74) | 1.02 (0.77, 1.36) |

| | | | | | | | |
|---|---|---|---|---|---|---|---|
| with prior coronary artery disease | >50 to ≤60 | 114,711 | 39,159 | 0.65 (0.42, 0.99) | 0.84 (0.74, 0.95) | 1.00 (0.74, 1.36) | 0.93 (0.81, 1.06) |
| | >60 to ≤70 | 142,095 | 61,631 | 0.81 (0.55, 1.19) | 0.89 (0.80, 1.00) | 1.04 (0.79, 1.37) | 0.96 (0.85, 1.09) |

**Table S7. Number of participants and coronary artery disease events (based on a secondary definition from previous MVP studies) for each MR design, with odds ratios and 95% confidence intervals corresponding to a 15.6 mg/dL increase in HDL-C or a 39 mg/dL increase in LDL-C obtained via one-sample (two-stage least squares) and two-sample approaches (inverse-variance weighting, MR-Egger regression, and weighted median regression)**

| Exposure and MR design | Number of participants | Number of events | Odds ratio (95% CI) | | | |
|---|---|---|---|---|---|---|
| | | | One-sample | Two-sample | | |
| | | | Two-stage least squares | Inverse-variance weighting | MR-Egger regression | Weighted median regression |
| **LDL (39 mg/dL increase)** | | | | | | |
| **Design 0**: Reference MR design | 366,492 | 68,668 | 1.49 (1.32, 1.66) | 1.14 (1.03, 1.28) | 1.36 (1.11, 1.66) | 1.26 (1.15, 1.38) |
| **Design 1**: Eligible participants aged 40-69 between Jan. 1, 2006 and Dec. 31, 2010 | 179,392 | 37,782 | 1.60 (1.36, 1.89) | 1.12 (1.00, 1.25) | 1.33 (1.08, 1.64) | 1.21 (1.09, 1.34) |
| **Design 2**: No adjustment for censoring (due to loss to or administrative end of follow-up) | 179,392 | 37,782 | 1.60 (1.35, 1.90) | 1.12 (1.00, 1.25) | 1.33 (1.07, 1.64) | 1.21 (1.09, 1.34) |
| **Design 3**: End of follow-up on March 31, 2017 without predefined length of follow-up | 179,392 | 39,198 | 1.63 (1.38, 1.93) | 1.15 (1.04, 1.28) | 1.35 (1.10, 1.64) | 1.27 (1.14, 1.41) |
| **Design 4**: Eligible participants included those with contraindication to LDL-C or HDL-C therapy | 193,960 | 42,903 | 1.59 (1.36, 1.86) | 1.15 (1.04, 1.27) | 1.34 (1.10, 1.63) | 1.28 (1.16, 1.42) |
| **Design 5**: Eligible participants included those with prior coronary artery disease | 235,829 | 84,772 | 2.19 (1.89, 2.54) | 1.29 (1.14, 1.45) | 1.57 (1.26, 1.96) | 1.47 (1.34, 1.60) |
| **HDL (15.6 mg/dL increase)** | | | | | | |
| **Design 0**: Reference MR design | 366,267 | 68,895 | 0.76 (0.68, 0.85) | 0.85 (0.76, 0.95) | 0.88 (0.66, 1.15) | 0.86 (0.77, 0.96) |
| **Design 1**: Eligible participants aged 40-69 between Jan. 1, 2006 and Dec. 31, 2010 | 179,631 | 38,036 | 0.81 (0.69, 0.94) | 0.88 (0.79, 0.97) | 1.00 (0.77, 1.29) | 0.90 (0.79, 1.02) |

| | | | | | | |
|---|---|---|---|---|---|---|
| **Design 2**: No adjustment for censoring (due to loss to or administrative end of follow-up) | 179,631 | 38,036 | 0.81 (0.69, 0.94) | 0.88 (0.79, 0.97) | 1.00 (0.78, 1.29) | 0.90 (0.79, 1.01) |
| **Design 3**: End of follow-up on March 31, 2017 without predefined length of follow-up | 179,631 | 39,291 | 0.82 (0.71, 0.95) | 0.89 (0.80, 0.99) | 1.07 (0.82, 1.39) | 0.94 (0.83, 1.06) |
| **Design 4**: Eligible participants included those with contraindication to LDL-C or HDL-C therapy | 194,190 | 42,998 | 0.99 (0.69, 1.43) | 0.88 (0.80, 0.98) | 1.02 (0.79, 1.31) | 0.94 (0.84, 1.06) |
| **Design 5**: Eligible participants included those with prior coronary artery disease | 236,020 | 84,828 | 0.80 (0.58, 1.09) | 0.87 (0.78, 0.97) | 1.03 (0.80, 1.34) | 0.95 (0.85, 1.05) |
